# Histopathological profile of triple negative breast carcinomas seen in patients in National Hospital, Abuja over a 10-year period (January 2010-December 2019)

**DOI:** 10.1101/2024.09.28.24314546

**Authors:** Tolulope A. Benye, Paul G. Jibrin, Ben I. Achusi, Friday G. Olah, Edmund J.C. Nwana

## Abstract

**Background:** Triple negative breast carcinoma (TNBC) cases in Africa and the black race which are most commonly seen among the reproductive age group are highly aggressive and have a high mortality rate. Based on its protein expression using immunohistochemical methods it does not express the hormone receptors (oestrogen and progesterone) and the human epidermal growth factor receptor 2 (HER2). As a result the only means of treating TNBC patients so far is by conventional chemotherapy +/- radiotherapy. Even then only 50.0 % of these patients will respond to the chemotherapy. Further studies which may help improve treatment and survival in these patients are important.

**Objective:** This study is aimed at knowing the histopathological profile of TNBCs in Abuja with the expression of androgen receptor; an important marker in these patients.

**Method:** Two hundred formalin-fixed paraffin embedded TNBC tissue blocks were selected for the study. The histological type and their respective histological grades (based on the Nottingham grading system) were noted from the haematoxylin and eosin(H&E) sections. Immunohistochemical staining was done using antibodies against androgen receptor (Biocare) and CK5/6 (Invitrogen) with in-house controls. The androgen receptor and CK5/6 expressions were analysed using a semi-quantitative scoring method: - A tumour was considered positive for LAR if ≥ 1.0 % of tumour cell nuclei were staining and positive for CK5/6 if ≥10.0 % of tumour cells had membrane staining. Other biographic data like tumour size, presence or absence of lymphovascular permeation were also analysed. Correlation between the histological types and grades were also done using SPSS version 21 statistical package.

**Results:** The 30-49 age group had the highest frequency of TNBC. The commonest histological type seen among TNBCs is Invasive carcinoma; NST at 93.0%. The most frequent histological grade is grade 2 at 56.0 %. The androgen receptor subtype constituted 19.0 %. The basal subtype formed 28.5 % of the total sample size. There was no significant correlation between the histological type and grade of TNBC. 73.3% of the patients had tumour size of pT1 and pT2.

**Conclusion:** This study shows that the percentage of androgen positive cases among triple negative breast carcinomas is significant enough to warrant routine luminal AR testing for TNBCs. Furthermore, this study has shown that the most frequent histological type is Invasive carcinoma; Non-specific type (NST) regardless of the molecular subtype and presence or absence of lympho-vascular permeation. It has also shown that the most frequent histological grade seen among patients with TNBCs is Grade 2.

## CHAPTER ONE

### 1.0 Introduction

Breast carcinoma is the second leading carcinoma in terms of incidence after carcinoma of the lungs globally. It is however, the most common cancer diagnosed in women and it constitutes about 24.2 % of all new cancer cases in women worldwide. In addition, it is also a leading cause of cancer-related mortality for women (15.0 %)^1^. In sub-Saharan Africa, breast carcinoma incidence (33.8 per 100,000 women per year) currently ranks second to cervical carcinoma incidence (34.8 per 100,000 women per year)^2^.

The prognosis of breast carcinoma is determined by histological type, staging, grade and molecular subtype. Current management guidelines are determined by the molecular subtype. Breast carcinoma has been historically sub-classified into four main molecular subtypes on the basis of three molecular markers; oestrogen receptor (ER), progesterone receptor (PR) and human epidermal growth factor receptor 2 (EGFR2/HER2)^3^.

These molecular subtypes include luminal type A; oestrogen receptor positive (ER+) and/or progesterone receptor positive [PR+], human epidermal growth factor 2 negative (HER2-), luminal type B (ER+ and/or PR+/-, HER2+) and HER2 type (ER-, PR-, and HER2+). The triple negative or basal subtype is negative for all these markers^4^.

In the latest College of American Pathologists/American Society of Clinical Oncology (CAP / ASCO) guidelines^5^, the threshold for HER2 positivity was reduced from 30.0 % to 10.0 % when using the immunohistochemical approach, and for in situ hybridization the HER2/CEP17 (HER2/ chromosome enumeration probe 17) ratio for HER2 gene amplification is now > 2 (reduced from >2.2), and a HER2 copy number of > 6 signals per cell is also sufficient for HER2 positivity. In addition, the threshold used for ER and PR positivity has been reduced to 1.0 % of the invasive cancer cells^6^. According to the 2017- 2018 Surveillance, Epidemiology and End Results (SEER) database, the majority (71.0 %) of breast carcinoma cases are hormone receptor positive, while 12.0 % are triple negative (TNBC)^7^. The TNBCs constitute 15.0 % to 20.0 % of breast carcinomas worldwide and they have unique characteristics^8,9^. The relative frequencies of TNBCs are higher in sub-Saharan Africa. It ranges from 28.0 % to 65.0 % in various studies done in Nigeria^10–12^. This has been attributed to several factors which include high parity, high frequency of obesity, poor health education, variations in pathology practices such as methods and duration of fixation, immunohistochemical methods and genetic influences^13^.

These TNBCs are seen in women of black race^14^. It is also the form of carcinoma with the worst prognosis due to the unique clinical and histopathological characteristics. These include younger age at presentation (< 50 years), high frequency in black women, high incidence of mutations in *BRCA1/2* genes, and having high-grade, high-stage, and high-proliferating tumours than those of western countries^9,15,16^.

The prognosis of triple negative breast carcinomas varies among individuals and has wide disparities in different parts of the world. This has led to many studies that have gradually shown through molecular analyses that it is a heterogeneous disease having a complex interaction between genes and environmental factors. This subsequently leads to variations in prognosis and treatment response^17^.

TNBCs have been characterised with high recurrence rate and poor response to conventional chemotherapy resulting in high mortality rates especially among the younger age group^18^.

TNBC is of great concern in Nigeria. The incidence has been shown to be on a steady increase^19^. This is worsened by poor health services, lack of expertise, financial constraints and illiteracy^20^.

In Nigeria, triple negative breast carcinomas have not been fully characterised. Different local studies have looked into a single type of classification among the different types of classification. This study will help to better characterise our TNBC patients into the different subtypes using positivity to androgen receptor and positivity/ negativity to CK5/6. This will help sub-classify these patients into the androgen receptor subtype and basal/ non-basal subtypes respectively. This will help understand this disease entity better in our own environment and also give a better outlook to our patients.

### 1.1 Statement of the problem

Triple negative breast carcinoma is a common disease in Nigeria with a poor response to treatment. It is common in pre-menopausal women and has a relatively high mortality rate. The class called TNBC in Nigeria is only diagnosed by exclusion i.e. by the negative outcome for immunostaining of oestrogen, progesterone and human epidermal growth factor receptors. It is treated as a single entity and subsequently managed with a singular treatment modality: - chemotherapy. Meanwhile, it is a heterogeneous disease with different sub-classes which can be determined by genetic studies and /or immunohistochemistry. As a result of this, prognosis is different among the subtypes. This sub-classification however is not yet routinely done in Nigeria.

### 1.2 Justification

The high incidence of TNBCs in our country and poor response to chemotherapy coupled with high mortality rates among the reproductive age group in this country makes this study of great importance. Paucity of data about the characteristics of TNBCs in Abuja and central parts of Nigeria e.g. Benue, Kogi, Kwara, Nasarawa, Niger and Plateau states in general, is one of the reasons for the motivation behind this study. The benefits of the androgen receptor subtype if TNBCs are accurately characterised as shown from different studies also proves this study of great importance.

### 1.3 Aim of the study

The aim of the study is to determine the histological characteristics and further characterisation of the triple negative breast carcinoma entity in patients seen in National Hospital, Abuja.

### 1.4 Objectives

1. To describe the age distribution and other biodata associated with triple negative breast carcinomas among patients in National Hospital.
2. To determine the histological type and tumour grade of TNBCs at presentation of patients seen in National Hospital and correlate between the histological type and tumour grade of triple negative breast carcinomas.
3. To determine the relative frequency of the luminal androgen receptor subtype of triple negative breast carcinomas seen in patients in National Hospital.

## CHAPTER TWO

### Literature Review

Breast carcinoma is the second most common carcinoma globally in women^1^. Breast carcinomas in Nigerian women tend to be of high-grade, high-stage, and high-proliferating subtypes occurring in a younger population than those of the Western countries^15^. Twenty-five to sixty-five percent of breast carcinoma cases in Nigeria are classified as triple negative. Although, it has been suggested that many of these cases are false negatives, resulting from inadequacies in tissue fixation and pathology practices. The mortality rates are high in our patients due to several factors which include poor access to health care, high poverty levels, poor enlightenment on breast cancer, traditional practices and beliefs, and relative lack of expertise and infrastructure^13^. Even though breast carcinomas are prevalent in this country, there are differences in their molecular and biological differentiation. These factors also vary among individuals within the same environment^18^.

An estimated one million cases of breast carcinomas are diagnosed annually worldwide^7^. Out of these, approximately 170,000 are of the triple-negative (ER–/PR–/HER2–) phenotype^7^. Triple negative breast carcinomas represent 10.0 % to 20.0 % of invasive breast carcinomas worldwide^21^. Out of these TNBC cases, about 75.0 % are basal-like^17^. Women with TNBC experience the peak risk of recurrence within 3 years of diagnosis, and the mortality rates are increased for 5 years after initial diagnosis. It has also been associated with black race, younger age at diagnosis, more advanced disease stage, higher grade, high mitotic indices, family history of breast carcinoma and *BRCA1* mutations^22^.

In a study done by Plasilova et al^23^ TNBCs were shown to be more common in women (13.0 %) than men (6.0 %). It was also shown that TNBC were highest under the age of 40 in whites, whereas in black patients, frequency did not reduce until after the age of 60. In the same study, it was shown that five-year survival is approximately 75.0 % for hormone receptors negative and triple negative breast carcinomas. This is lower than that of hormone receptor and triple positive breast carcinomas (90.0 %). It has also been shown that the frequency rates for breast carcinoma death is approximately 7.5 % per year. This is for the first two years following diagnosis for hormone receptor negative and triple negative tumours. Thereafter, the rate declines rapidly. In contrast, hormone receptor positive tumours have frequency rates that lack a sharp peak but are relatively constant at 1.0 % to 2.0 % per year^23^.

Mutations in tumour suppressor genes such as *BRCA1* have been implicated in the higher TNBC frequencies in Nigeria. A multi-centre study by Wright et al^13^ has shown that there are multiple biomarkers that are generally overexpressed in Nigerian and West African patients with breast carcinoma, are associated with TNBC status and aggressive clinical and pathological characteristics^13^. In this study also, there is an International Consortium for Advancing Research on TNBC strategic action plan for Nigeria due to the burden of the disease^13^. This is to help prompt diagnosis, treatment and enable further research into this disease entity as is unique to our biology.

The relative frequencies of TNBCs differ in different parts of the world. About 10.0 % to 20.0 % of invasive breast tumours belong to the triple-negative category worldwide^22^. In Nigeria, different studies have shown higher frequencies than what obtains worldwide; 28.0 % to 65.0 % of invasive breast carcinomas^10,11^. It has been postulated that this wide variation could be as a result of variations in sample size and pathology practices^13^. It has been shown in West Africa (which is inclusive of some countries in Sub-Saharan Africa), that high frequencies are observed with values ranging from 27.0 % to 61.0 %. In North Africa also, the range is 17.0 % to 29.0 %. This is similar to findings seen in other parts of Africa. Even though the frequencies are high, countries in North Africa have relatively lower values than other African countries^24^. In Asia, frequencies are also high varying between 20.5 % to 36.7 % in different regions^25^. Frequencies are lower in Europe^26^ and in North America^24^. They are however higher in some parts of South America^27^. Hormone receptor negative breast carcinomas inclusive of triple negative breast carcinomas are associated with early-onset carcinomas (earlier than 40 years of age) whereas hormone receptor positive carcinomas are associated with late-onset disease (later than 50 years of age)^28^.

The Nottingham (Elston-Ellis) modification of the Scarff-Bloom-Richardson grading system, also known as the Nottingham Grading System (NGS), is the grading system recommended by various professional bodies internationally (World Health Organization [WHO], American Joint Committee on Cancer [AJCC], the National Health Scheme(NHS), UK and the Royal College of Pathologists (UK RCPath) to assess histological grade and by extension, prognosis^29^. The NGS is based on the evaluation of three morphological features: (a) degree of tubule or gland formation, (b) nuclear pleomorphism, and (c) mitotic count. Each is scored from 1 to 3. Adding the scores gives the overall histological grade. This is shown in Appendix 5.

In Nigeria, majority of the patients (up to 60.0%) are ≤ 50 years of age at diagnosis^13^ and tumour grade was variable between the regions. However, Grades 2 and 3 tumours have the highest frequencies^10^. This finding is also similar to what is observed in sub-Saharan Africa, North Africa, Europe^26,30^, Asia^25,31^, and the Americas^24,32^. Even though the frequencies of Grades 2 and 3 tumours are higher in Russia in a study conducted by Zhukova^30^, these tumour grades are slightly lower in other parts of the world when compared to Nigeria.

Traditionally, breast carcinomas including triple negative breast carcinomas have been classified based on morphological and histological characteristics. Based on this, there are different types of invasive carcinomas of the breast. It has been shown that a high proportion (74.0 % to 83.0 %) of TNBCs are diagnosed as invasive ductal carcinoma, not otherwise specified (NOS) morphology. Other variants of TNBC include medullary carcinoma, apocrine carcinoma, adenoid cystic carcinoma, carcinomas arising in microglandular adenosis, myoepithelial carcinomas, and metaplastic carcinomas^23^. Some studies in Nigeria have shown that the TNBCs were predominantly invasive ductal carcinomas; No Special Type (NST) constituting 90.7 % to 92.3 % of cases. Other histological types reported include invasive lobular, mucinous, invasive papillary carcinomas and invasive ductal carcinoma with squamoid differentiation^12,33,34^. In Egypt and Eastern Algeria (North Africa), invasive ductal carcinoma; NOS type is also the most common^24^. This is similar to findings observed in Asia, Europe and the Americas^24–26^.

The molecular classifications of TNBCs were introduced to determine the prognosis and outcome in patients with this disease. There are different classifications, each using specific parameters. These include the transcriptome, genome, epigenome, proteome, relationship to *BRCA* and immune profile bases of classification^35^. Each of the different modes of classification has its unique characteristics. There are however two broad classifications of TNBC basically; the basal type and non-basal type depending on the expression of the basal myo-epithelium marker signified by cytokeratin 5/6 (CK5/6). To a large extent, all TNBCs are generally classified molecularly as basal type. This is because the basal type constitutes 50.0 % to 75.0 % of TNBCs and these types can be identified by immunohistochemistry^17^. The basal type tumours are often referred to as TNBCs because majority of basal-like tumours are typically negative for oestrogen receptor (ER), progesterone receptor (PR), and human epidermal growth factor receptor (HER2). These tumours are Retinoblastoma gene (*Rb*) and *p53* deficient and so, they are likely to grow rapidly. This subtype of breast carcinoma is characterized by a gene expression profile that is similar to that of the basal myo-epithelial layer of the normal breast due to its expression of cytokeratin 5, 6 or 17. Genes associated with cell proliferation, such as marker of proliferation *Ki67* and topoisomerase (DNA) II alpha (*TOP2A*), are also highly expressed in basal-like tumours^17^.

Several modifications have been made to the classification of the TNBCs using the transcriptome as the basis for classification. According to Lehmann’s classification, TNBC molecular subtypes have been modified from six (TNBC type) into four (TNBCtype-4) tumour-specific subtypes. These include Basal-like type 1 (BL1), Basal-like type 2 (BL2), Mesenchymal (M) and Luminal Androgen receptor (LAR). This classification demonstrates differences in diagnosis, age, tumour grade, local and distant disease progression^36^. Progressive modifications of its molecular classification are as shown in Appendix 3. The different parameters used in sub-classifying TNBCs^35^ are as shown in Appendix 4. Expression of either CK5/6 or epidermal growth factor receptor (EGFR) has been shown to accurately identify basal-like tumours classified using gene expression. Approximately 75.0 % of TNBCs are basal-like, with the other 25.0 % comprising all other mRNA subtypes^37^. In a study by Titloye et al^38^ in Ife, Nigeria however, the predominant molecular phenotype was the non-basal, triple-negative type (47.65%). This non-basal group does not express basal cytokeratins but rather express the luminal forms which include CK 8/18 or 19. These subtypes include mostly HER2-positive breast carcinoma and also breast carcinomas that lack ER, PR, and HER2 in immunohistochemical analysis but do not exhibit the features of the basal- like subtype. The potential therefore exists for misclassification if basal-like biology and triple negative phenotype are used to identify patients^17^. Among newly diagnosed breast carcinoma patients also, < 10.0 % have *BRCA1* or *BRCA2* mutated genes but this percentage is higher among patients with TNBC with around 35.0 % of *BRCA1* and 8.0 % of *BRCA2* mutations in this population. Among *BRCA1* mutation carriers, more than one- third have TNBC^17^.

Masuda et al^39^ confirmed the six molecular subtypes of triple negative breast carcinomas as advanced by Lehmann. The six TNBC subtypes include two basal-like (BL1 and BL2), one immune-modulatory (IM), one mesenchymal (M), one mesenchymal stem-like (MSL), and one luminal androgen receptor (LAR) subtype, the last being characterized by androgen receptor (AR) signalling. Burstein et al^40^ were further able to revise the Lehmann’s classification and distinguish four stable, molecularly-defined TNBC subtypes: luminal androgen receptor (LAR), mesenchymal (MES), basal-like immune-suppressed (BLIS), and basal-like immune-activated (BLIA). MES, BLIS, and BLIA are characterized by distinct clinical prognoses, with BLIS tumours having the worst and BLIA tumours having the best outcome. A recent study by Lehmann^36^ reported that the previously described Immune-modulatory and Mesenchymal stem-like subtypes were contributed from infiltrating lymphocytes and tumour-associated stromal cells respectively. This refined TNBC molecular subtypes from six (TNBCtype) into four (TNBCtype-4) tumour-specific subtypes (BL1, BL2, M, and LAR).

There are different characteristics of each of the molecular types of TNBC. The basal-like subtypes (BL1 and BL2) are characterized by enrichment of cell cycle and cell division components and pathways, elevated DNA damage response (*ATR/BRCA*) pathways, growth factor signalling pathways (Epidermal growth factor; EGF, Nerve growth factor; NGF, MET, *Wingless/Beta-Catenin; Wnt/β-catenin*), and Insulin-like growth factor 1 receptor; IGF1R as well as glycolysis and gluconeogenesis. The immune-modulatory subtype constitutes approximately 20.0 % of TNBCs and are highly enriched in immune cell markers and signalling^41^. Tumours that have more than 50.0 % lymphocytic infiltrates are considered lymphocyte-predominant breast carcinoma and have the best prognosis. The mesenchymal and mesenchymal stem-like (M and MSL) subtypes are heavily enriched in components and pathways involved in cell motility, extracellular receptor interaction and cell differentiation pathways. They also express genes representing components and processes linked to growth factor signalling pathways that include inositol phosphate metabolism, Epidermal Growth factor receptor (EGFR), Platelet-derived Growth factor (PDGF), calcium signalling, G-protein coupled receptor, and Extracellular signal-regulated kinase (ERK1/2) signalling as well as ATP-binding Cassette (ABC) transporter and adipocytokine signalling^40^. All the earlier stated subtypes (the immune-modulatory, the mesenchymal and the mesenchymal stem-like) need genomic profiling to accurately characterize them^39^.

The Luminal androgen receptor subtype is the most unique among the TNBC subtypes. This subtype is oestrogen receptor negative (ER -), but gene signatures are heavily enriched in hormonally regulated pathways which include steroid synthesis, porphyrin metabolism, and androgen/oestrogen metabolism. This subtype of tumours exhibit androgen receptor, oestrogen receptor, prolactin, and ERB-B2 Receptor Tyrosine Kinase 4 (ErbB4) signalling, but oestrogen receptor-α (ER-alpha) negative immunohistochemical staining. However, these ‘ER-negative’ tumours demonstrate molecular evidence of ER activation. This may be because less than 1.0 % of these tumour cells express low levels of ER protein, defining them as ‘ER-negative’ by immunohistochemistry. Tumours within the LAR group express numerous downstream androgen receptor (AR) targets and co-activators as well. Androgen receptor (AR) expression by immunohistochemistry in the LAR subtype is more than 10-fold higher compared to other TNBC subtypes^17^.

The androgen receptor has a complex interaction with the oestrogen receptor making treatment modalities variable^41^. In the presence of the hormone receptors ER and PR, the AR serves as an antagonist to their growth-promoting effects whereas in their absence, it can singularly drive growth stimulating effects. This results in variations in therapeutic regimen for this group of patients. The androgen status in TNBCs has become important as both a prognostic marker and potential therapeutic target in breast carcinomas. Androgen receptor (AR) expression by immunohistochemistry (IHC) has been observed in 90.0 % of primary breast carcinomas and in 75.0 % of breast carcinoma metastases^41^. The highest positivity is observed in oestrogen receptor (ER)-positive tumours (80.0 % to 90.0 %) of cases and the lowest is seen in triple-negative tumours (30.0 %). Although AR can be expressed in multiple molecular subtypes of TNBC, the luminal androgen receptor (LAR) subtype has the highest level of AR expression. In ER positive breast carcinoma, AR signalling has been correlated with a better prognosis because of its inhibitory activity in oestrogen dependent disease^41^.

The LAR subtype is predominantly sub-classified in the non-basal subgroup and represents a subtype of TNBC with a distinct prognosis^41^. The frequency of AR expression in TNBC has a wide range of 6.6 % to 75.0 % in literature. This heterogeneity is as a result of various reasons which include the number of patients in the study and the cut-off used for AR positivity (≥1.0 % or ≥10.0 %), the source of the primary antibody, the methodology of testing and the effects of patient selection in prospective studies^41^.

The androgen signalling pathway plays a critical role in the development of normal and malignant breast tissue. Androgens such as testosterone and dihydrotestosterone (DHT) can have either an inhibitory or a stimulatory effect on breast carcinoma cell lines depending on the co-expression of other steroid hormone receptors and the presence or absence of breast adipose tissue fibroblasts (BAFs)^42^. The androgen receptor (AR) has been shown to be a marker of low-grade, well-differentiated disease. In TNBC tumours, AR positivity by immunohistochemical staining is a favourable prognostic factor. It is also associated with a lower clinical stage, lower histologic grade, and lower mitotic score. It has also shown that the absence of AR expression is associated with an increased risk for recurrence and distant metastasis in lymph node positive TNBCs^42^.

Data on this subtype from indigenous studies in Nigeria is not readily available however, a multi-institutional cohort study showed wide variations in the expression of the androgen receptor in TNBCs. This study showed that the United Kingdom, Norway, Ireland, United States, India and Nigeria had percentage expression of the androgen receptor of TNBC samples being at 54.9 %, 32.7 %, 24.7 %, 24.8 %, 22.7 % and 8.3 % respectively. Nigeria had the lowest expression among other countries^43^. Also, in comparison to other countries in Africa with frequencies of 44.0 % (Ghana) and 60.7 % (Egypt)^44^, Nigeria still has the lowest however, higher levels of expression have been reported in Asia, Europe and the Americas^43^.

## CHAPTER THREE

### Materials and methods

#### 3.1 Study design

This study is a descriptive retrospective study of all female breast carcinoma specimen diagnosed as triple negative shown through negativity for Oestrogen, progesterone and Human epidermal growth factor markers received in the Department of Histopathology, National Hospital between the 1st of January 2010 to 31st of December 2019.

#### 3.2 Study site and population

National Hospital is a tertiary hospital with established oncology services. It is situated in the city of Abuja, Federal Capital Territory (FCT) which has a population of 3.3 million^45^. It also serves as referral centre for oncology services to other tertiary institutions within FCT and beyond.

The Department of Histopathology in National hospital is equipped with semi-automated and fully automated histopathology and immunohistochemistry services.

This work was conducted on all formalin-fixed, paraffin-embedded blocks of already confirmed triple negative female breast carcinoma samples at the Histopathology department of National Hospital, Abuja which met the inclusion criteria over the study period.

The average number of surgical samples received in the department is about two thousand cases per year. An average of two hundred and sixty cases are breast samples (benign and malignant) and an average of one hundred and twenty samples are diagnosed as breast cancer received yearly.

#### 3.3 Sample size determination

Convenience sampling of readily available two hundred and ten triple negative formalin-fixed, paraffin-embedded tissue blocks was done. Two hundred samples met the inclusion criteria.

#### 3.4 Sampling method

All triple negative formalin-fixed, paraffin-embedded blocks were retrieved using the department’s database for the study period. Demographic data and tumour characteristics were obtained from the database using a standard protocol. Tumour grade was assessed according to the modified Nottingham Scarff-Bloom-Richardson grading system^29^ as shown in appendix 5.

#### 3.5 Inclusion criteria

1. All triple negative female breast carcinoma samples received within the study period
2. Slides with complete and adequate paraffin blocks
3. Slides with representative standard staining required for diagnosis
4. Samples with complete records
5. Patients that have more than one biopsy specimen had just one of the samples regarded as study sample for the patient. This was done by merging the data of all the years of study involved. Thereafter, the conditional formatting tool in the Microsoft excel software was used to highlight each of the names and/ or hospital numbers for each patient starting from the first year of study. If there are similar multiples of such patient, the other samples are deleted from the database being used for study (inclusive of the samples on which immunohistochemistry was not done at all). A lot of the patients in this study had immunohistochemistry done just once making it easy for elimination and data analysis.

#### 3.6 Exclusion criteria

1. Slides that do not have tumour
2. Metastatic breast carcinoma and /or carcinoma of non-breast origin were excluded.
3. Cases of pure ductal carcinoma in-situ without invasion were excluded.
4. Slides with missing or distorted paraffin blocks were excluded.
5. All male breast carcinoma samples were excluded.
6. All samples with a HER-2 ASCO/CAP score of +2 (equivocal) were excluded.
7. Samples on which immunohistochemistry was not done at all.
8. Other breast malignancies were excluded.

Three slide sections were obtained from the formalin-fixed paraffin embedded tissue blocks of samples that met the inclusion criteria. Blocks were sectioned at 4 micrometers for optimal quality. Out of the three sections, one was stained with haematoxylin and eosin for assessment of histological type and grade. The other two slides were used for immunohistochemistry. For all the triple negative breast carcinoma samples, all haematoxylin and eosin–stained slides were reviewed and the original histological details confirmed, without prior knowledge of patient demographics. The histological subtypes were re-classified using the WHO classification of breast tumours; 4^th^ edition ^47^ as shown in appendix 8. Tumours were graded based on the modified Nottingham Scarff-Bloom-Richardson grading system. Mitotic counts were performed over ten high power fields per slide^29^.

#### 3.70 Immunohistochemistry

All samples which met the inclusion criteria were subjected to immunohistochemistry (IHC analysis). Immunohistochemical analysis was performed using androgen receptor and Cytokeratin 5/6 monoclonal antibodies and representative blocks selected per case for analysis. Details of the antibodies, their clones, dilutions and sources are provided as in Appendix 9.

#### 3.71 Classification based on immunohistochemistry

Tissue blocks with nuclear staining in ≥ 1.0% of tumour cell nuclei for the androgen receptor antibody were classified as androgen receptor positive and labelled as the androgen receptor subtype. Tissue blocks with membrane staining in ≥ 10.0% of tumour cell nuclei for the cytokeratin 5/6 (CK5/6) antibody were classified as CK5/6 positive and labelled as basal subtype and those not staining for CK5/6 were classified as the non-basal subtype. The non-basal subtype were further sub-classified into the androgen receptor subtype and the non-androgen receptor subtype i.e. the mesenchymal subtype.

#### 3.8. Immunohistochemistry protocol

The formalin-fixed, paraffin-embedded triple negative breast carcinoma tissue blocks were used in this study and the interpretation of the molecular tests done (androgen receptor and CK5/6) are as follows; Basal type (Androgen receptor negative and Cytokeratin 5/6 positive), non-basal type (Cytokeratin 5/6 negative with a positive or negative test for androgen receptor) and Androgen receptor type (defined as positivity for androgen receptor only). Cut off values of ≥ 1.0 % and ≥10.0 % were applied for the androgen receptor and cytokeratin 5/6 scoring respectively (Appendix X).

Tissue specimens and clinical information were analysed.

#### 3.90 Data analysis

Data analysis was performed using the Statistical Package for Social Sciences (SPSS Inc., Chicago, IL, USA) version 21 and Microsoft Excel version 16. Descriptive analysis was performed to characterize the demographic variables of the patients. Mean and standard deviation (SD) were determined for the continuous variables with normal distribution and ranges for the continuous variables with skewed distribution.

Frequency tables, pie charts and bar charts were used to present data on the age of patients and relative frequencies of TNBCs, relative frequencies of histological type and grade, androgen receptor subtypes and that of basal and non-basal types.

Statistical comparisons between groups were completed using the Pearson’s chi-square test as appropriate.

Results of the molecular testing were analysed based on histological type and histological grade. P < 0.05 was considered statistically significant.

#### 3.10 Limitations to the study

1. The manner of processing of the tissue blocks and the pre-analytical conditions of the tissue such as cold ischaemic time, quality of fixation and processing especially for those from referral centres could not be influenced since the samples were archival.
2. Exclusion of samples with HER-2 scores of +1 and +2, not being further evaluated by Fluorescence in-situ hybridization (FISH), may have affected the sample size of those being classified as triple negative and subsequently the samples available for the androgen receptor and CK5/6 testing.

#### 3.11 Ethical considerations

Ethical approval was obtained from the National Hospital Ethical Review Committee (Appendix XI).

#### 3.12 Confidentiality statement

There was strict adherence to patients’ confidentiality and their identities were always concealed. The specimens were not used for any other purpose aside from what was mentioned in this research work. No further consent was sought other than the one implied by submission of samples. This is because no genetic tests were done at this stage.

## CHAPTER FOUR

### Results

During the study period (January 2010 to December 2019), a total number of 3,389 breast samples were received. One thousand six hundred and eighty-eight (n=1,688) of them were invasive breast carcinomas. Only one thousand and forty-six of them (n=1,046) met the inclusion criteria.

The triple negative breast carcinomas constituted 19.12 % (n=200) of the invasive carcinomas analysed. The other molecular subtypes seen in this study were as follows; the luminal type A (42.92 %), luminal type B (18.07 %); (total – 60.99 %) and the Her-2 type (19.89 %).

From the triple negative carcinoma cases, right sided carcinoma cases constitute 50.50% (n=101) and the left sided cases constitute 49.50% (n=99). There were no bilateral cases seen.

The TNBCs were categorized into basal subtypes based on positivity to cytokeratin 5/6 staining (shown as membrane staining seen in ≥ 10.0% of tumour cells in a sample) and non-basal subtypes based on negativity to cytokeratin 5/6 staining (shown as membrane staining seen in ≤ 10.0% of tumour cells in a sample). The non-basal type constitutes 71.50 % of the TNBCs (n=143/200) while the basal type constitutes 28.50 % in this study (n=57/200 as shown in Table III). The non-basal type is further categorized into the androgen receptor and non-androgen receptor subtypes i.e. mesenchymal subtypes (Table IV). The androgen receptor subtype constitutes 19.0 % of the TNBCs (n= 38/200 as shown in Table II) and the mesenchymal subtype constitute 52.50 % (Fig 3).

The most common histological type among all the invasive breast and triple negative breast carcinomas is Invasive carcinoma; Non-Specific Type (NST) constituting 95.79 % (Table VI) and 93.0 % (Table VIII) respectively. The most common histological grade is Grade 2; also classified as low-grade tumours constituting 60.90 % (Table VII) and 56.0 % (Table VIII) respectively. This is followed by grade 3; also classified as high-grade tumours constituting 23.0% and 9.8% respectively.

There is no correlation between the histological types and histological grades of TNBCs.

Tumour sizes could be determined for 52.5% of the samples (n=105). A large number had tumour sizes of < 2.0cm (n=41/105) followed by those of tumour sizes ≥2.0cm but less than 5.0cm (n=36/105). Lympho- vascular permeation could be determined in 18.0% of the total sample size (n=36/200) as shown in Table X).

#### 4.1 Age distribution and relative frequency of triple negative breast carcinomas among invasive breast carcinomas seen in patients in National Hospital

**Table I:**
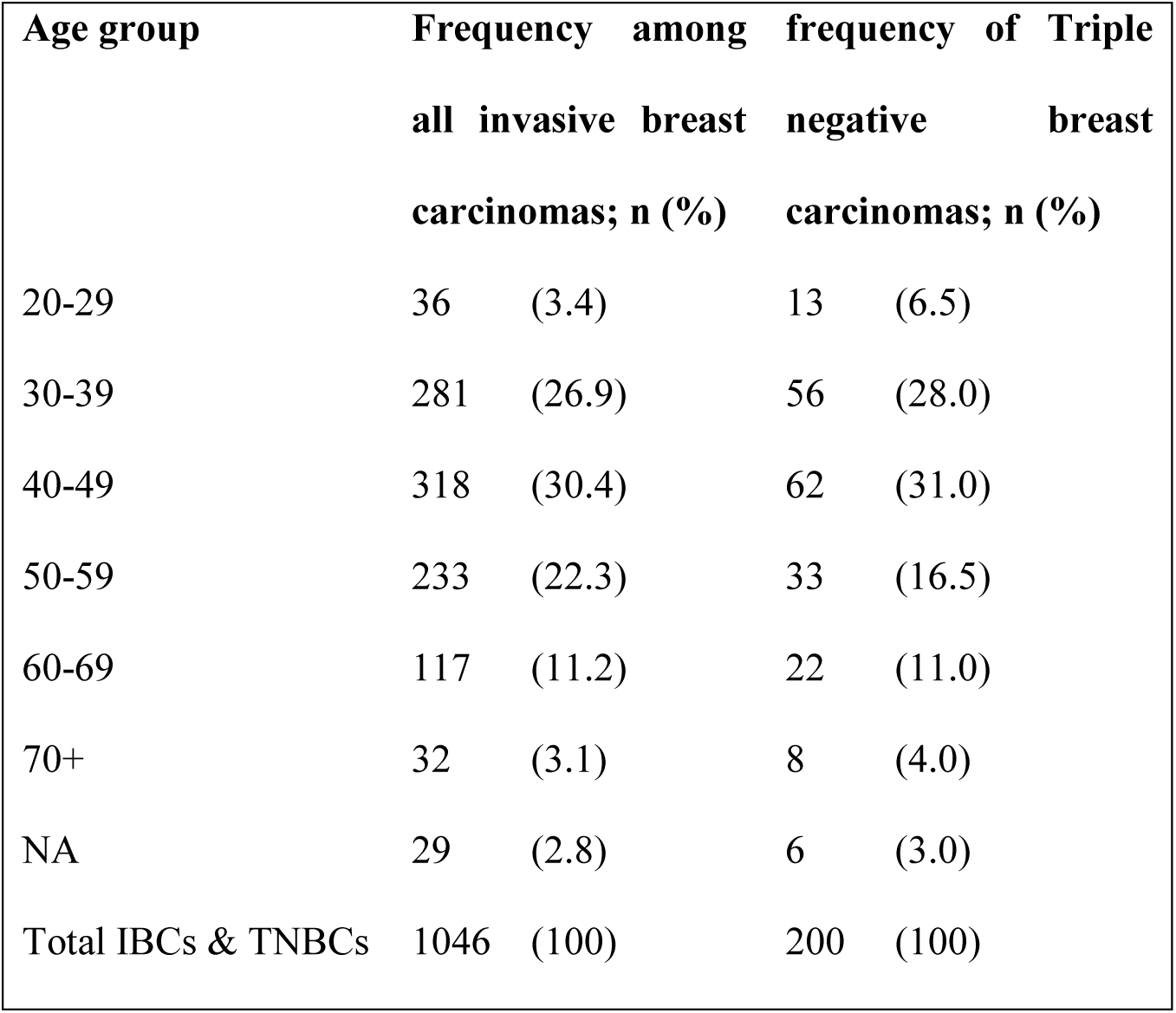
Age distribution and relative frequency of patients with triple negative breast carcinomas among invasive breast carcinomas.

Table I above shows the highest frequency of TNBCs is found among the premenopausal age group (30- 49).

**Figure 1:**
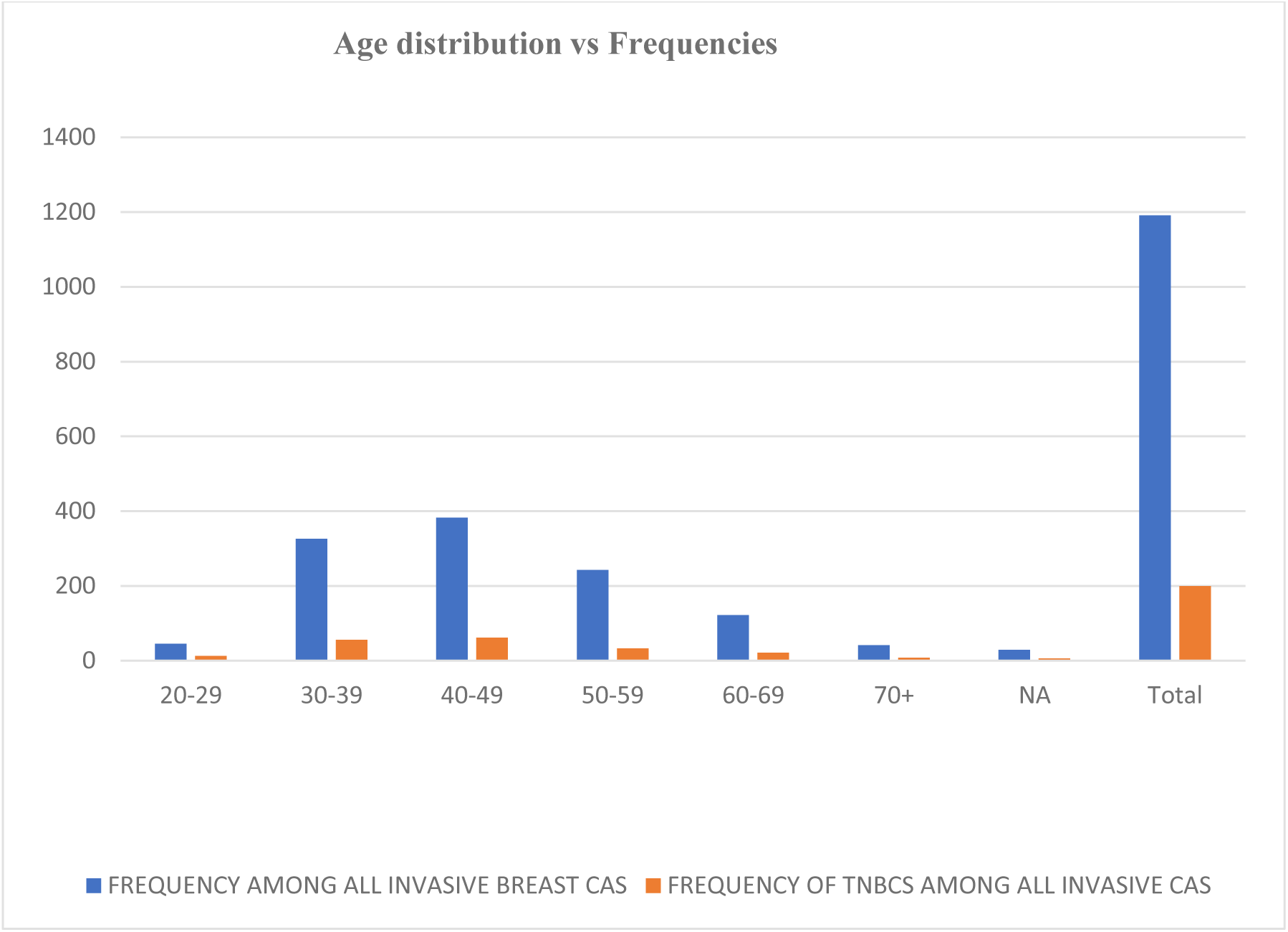
A clustered column chart comparing the relative frequencies of the different age groups as seen in all invasive carcinomas and triple negative carcinomas.

#### 4.2 The relative frequency of the Androgen receptor subtype of triple negative breast carcinomas seen in patients in National Hospital

**Table 2:** Distribution of the Androgen Receptor subtype of triple negative breast carcinomas.

| **AR status** | **Frequency** | **Percentage** |
| --- | --- | --- |
| Negative | 162 | 81.0 |
| Positive | 38 | 19.0 |
| **Total** | **200** | **100.0** |

The table above shows that 19.0 % of breast carcinomas are of the luminal androgen subtype.

**Figure 2:**
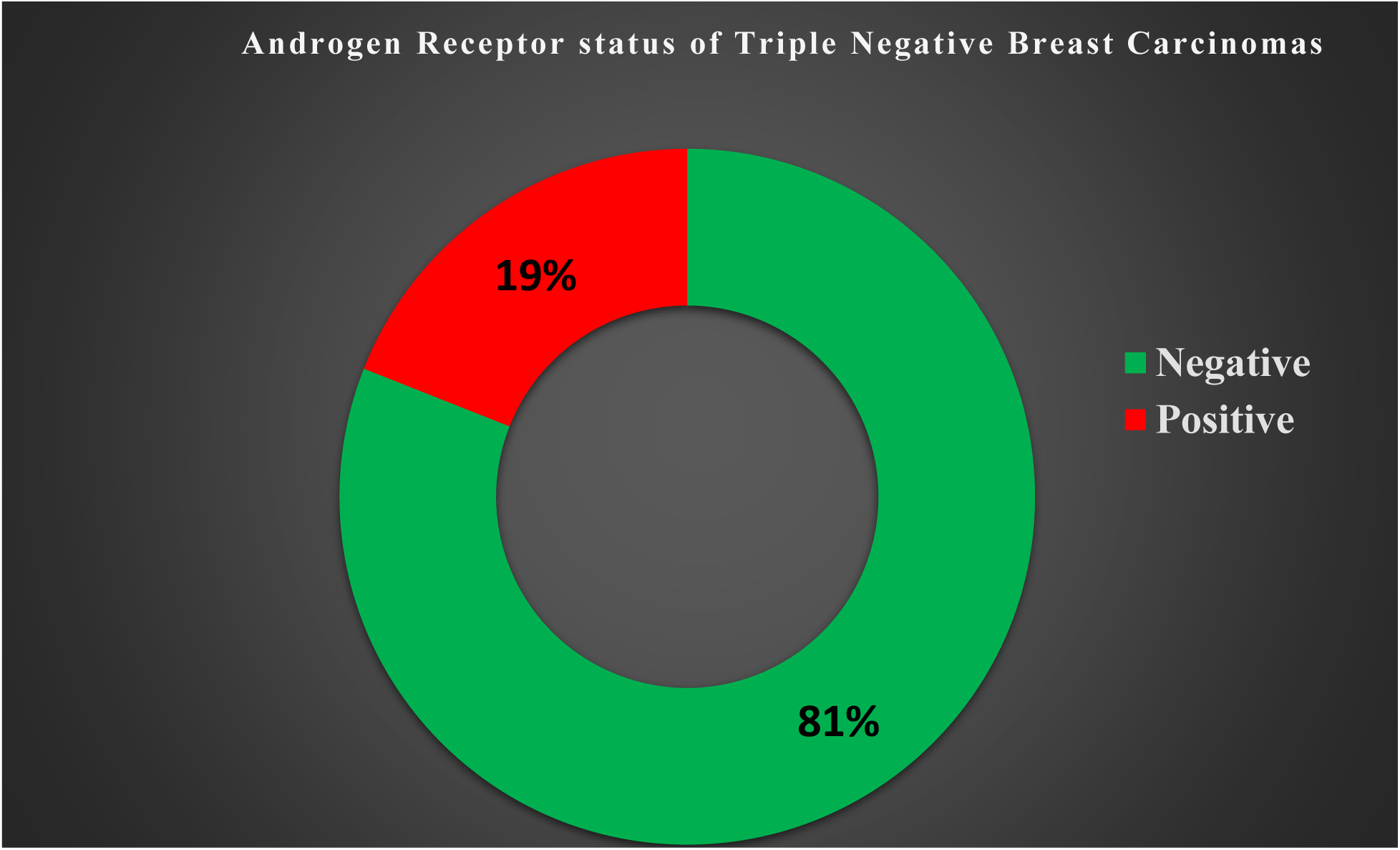
A pie chart showing the frequency of androgen receptor status.

The above figure illustrates that 19.0 % of TNBCs are positive for androgen receptor.

**Table 3:** Distribution of cytokeratin 5/6 (CK5/6) status among patients with the triple negative breast carcinomas.

| **Cytokeratin 5/6 status** | **Frequency** | **Relative frequency (%)** |
| --- | --- | --- |
| Basal type (positive) | 57 | 28.5 |
| Non-Basal type (Negative) | 143 | 71.5 |
| Total | 200 | 100 |

The table above shows the non-basal type has the highest frequency with a percentage of 71.50 % (143/200)

**Table 4:** Combined frequencies of the non-basal type among patients with triple negative breast carcinoma.

| **CK5/6 Negative** | **Frequency** | **Relative frequency (%)** |
| --- | --- | --- |
| (AR status) Positive | 38 | 19.0 |
| (AR status) Negative | 105 | 52.5 |
| Total | 143 | 71.5 |

The table above shows the samples that are both CK5/6 and AR negative have the highest frequency with a percentage of 52.5 % (105/143) while those that are CK5/6 negative but AR positive (androgen receptor subtype) have a percentage of 19.0% (38/143).

**Figure 3:**
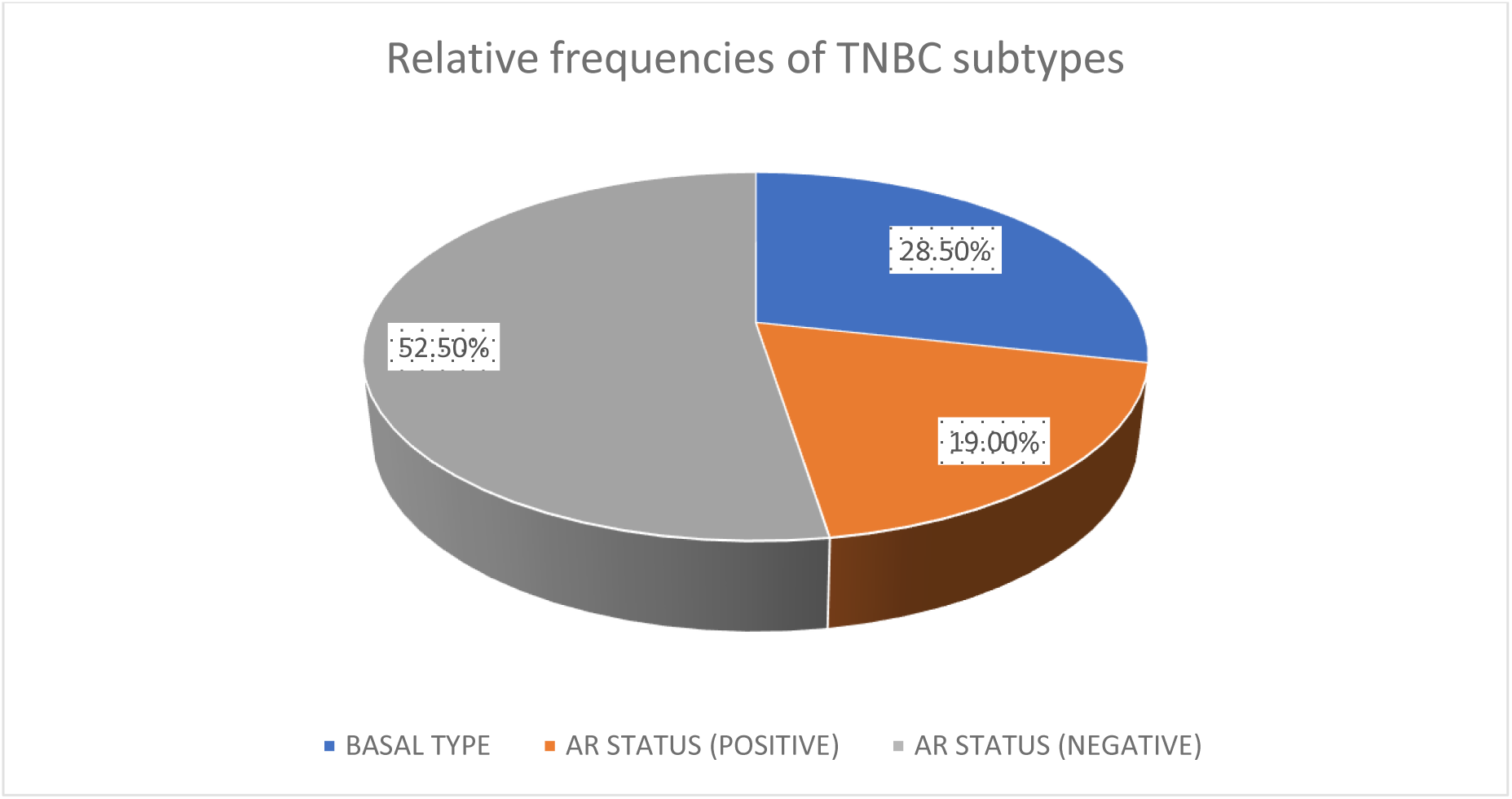
A pie chart showing the frequencies of the TNBC molecular subtypes with the mesenchymal subtype being the most common constituting 52.5 %.

#### 4.22 Age group-matched frequencies of the androgen receptor subtype

**Table 5:** Age-group matched frequencies of androgen receptor subtype.

| AGE GROUP | FREQUENCY | (%) TOTAL |
| --- | --- | --- |
| 20-29 | 1 | 2.6 |
| 30-39 | 8 | 21.1 |
| 40-49 | 15 | 39.5 |
| 50-59 | 6 | 15.8 |
| 60-69 | 5 | 13.2 |
| s≥70 | 2 | 5.3 |
| ADULT | 1 | 2.6 |
| TOTAL | 38 | 100.00 |

The table above shows the premenopausal age group (30-49) with the highest frequency of the AR subtype

#### 4.30 Distribution of the histological types seen among patients with invasive breast carcinomas

**Table VI:** Relative frequencies of the different histological types of all invasive breast carcinomas.

| **Histological Type** | **Frequency** | **% Total** |
| --- | --- | --- |
| Invasive carcinoma; NST | 1002 | 95.8 |
| Carcinoma with apocrine differentiation | 1 | 0.1 |
| Inflammatory Carcinoma | 1 | 0.1 |
| Invasive Lobular Carcinoma | 9 | 0.9 |
| Invasive Papillary Carcinoma | 5 | 0.5 |
| Invasive Tubular Carcinoma | 2 | 0.2 |
| Carcinoma with medullary features | 10 | 1.0 |
| Metaplastic Carcinoma | 4 | 0.4 |
| Mucinous Carcinoma | 12 | 1.2 |
| **Total** | **1046** | **100.2** |

This table shows Invasive carcinoma; NST as the most common histological type constituting 95.8 % (1002/1046).

#### 4.31. Distribution of histological grade among patients with invasive breast carcinomas

**Table VII:**
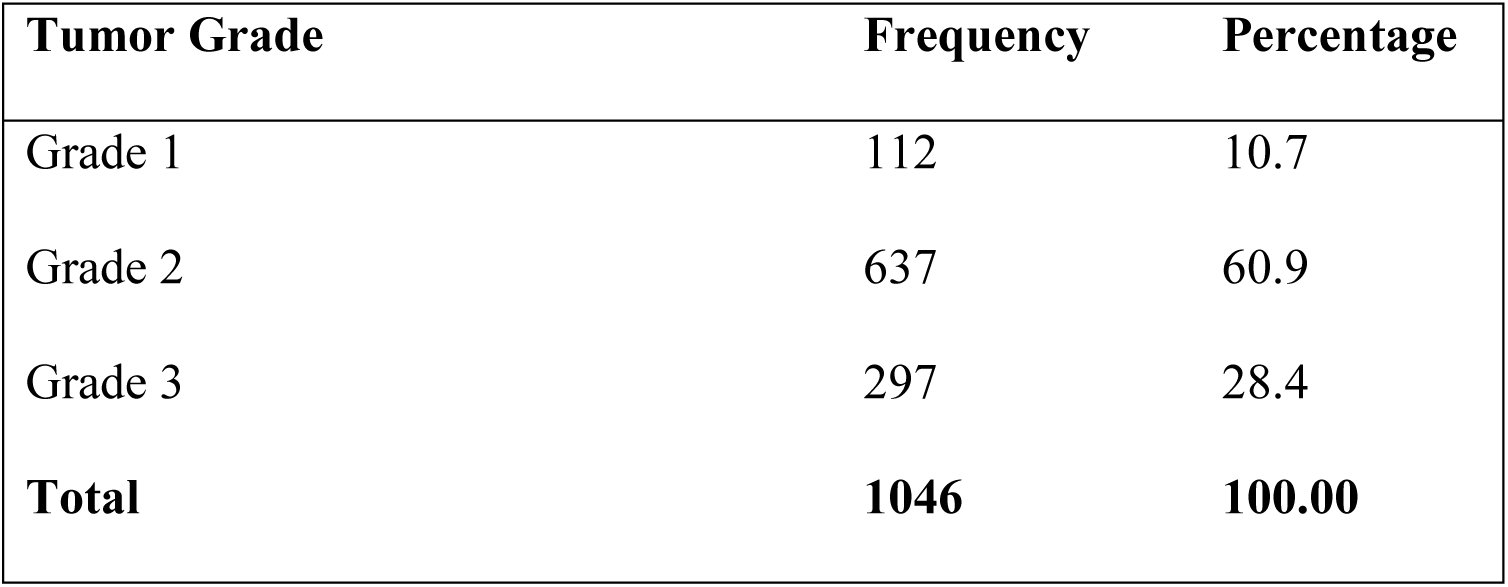
Relative frequencies of the different histological grades of all invasive breast carcinomas.

| **Tumor Grade** | **Frequency** | **Percentage** |
| --- | --- | --- |
| Grade 1 | 112 | 10.7 |
| Grade 2 | 637 | 60.9 |
| Grade 3 | 297 | 28.4 |
| **Total** | **1046** | **100.00** |

The table above shows that grade 2 has the highest frequency at 60.9 % (637/1046)

#### 4.4. The correlation of histological types and grades of triple negative breast carcinomas

**Table VIII:**
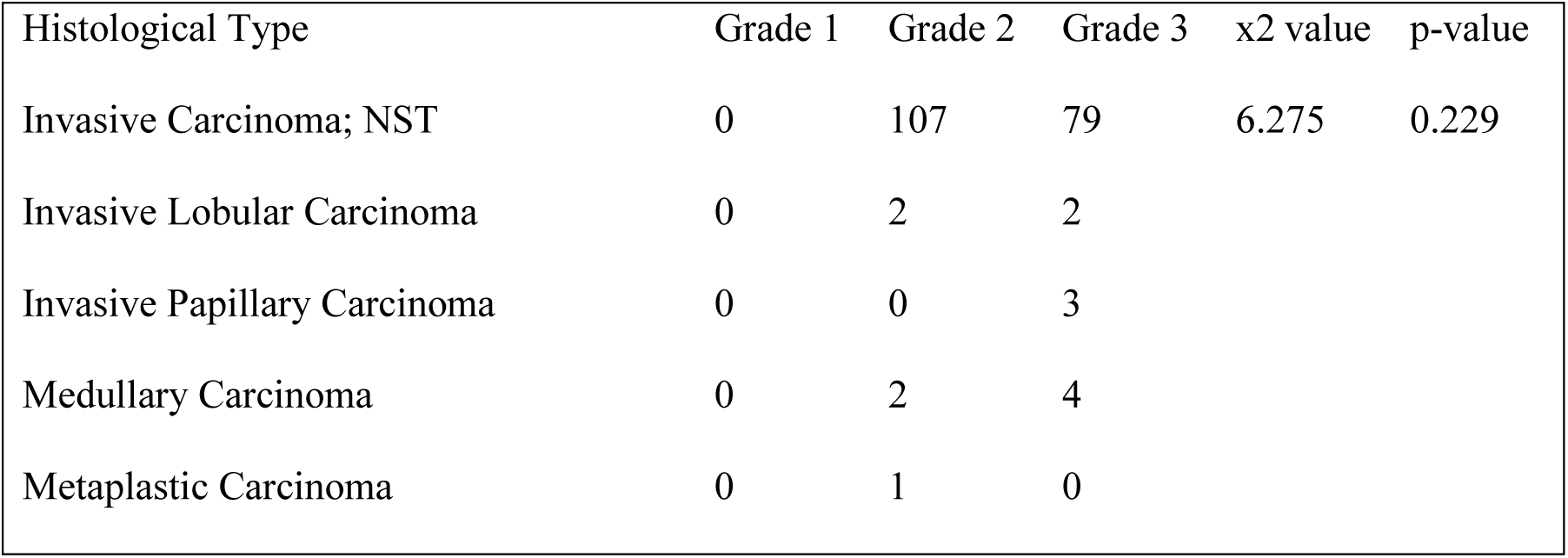
The frequencies of the different histological types and histological grades among the Triple negative breast carcinomas.

| Histological Type | Grade 1 | Grade 2 | Grade 3 | x2 value | p-value |
| --- | --- | --- | --- | --- | --- |
| Invasive Carcinoma; NST | 0 | 107 | 79 | 6.275 | 0.229 |
| Invasive Lobular Carcinoma | 0 | 2 | 2 |  |  |
| Invasive Papillary Carcinoma | 0 | 0 | 3 |  |  |
| Medullary Carcinoma | 0 | 2 | 4 |  |  |
| Metaplastic Carcinoma | 0 | 1 | 0 |  |  |

The table above shows that invasive carcinoma; NST histological type has the highest frequency (186/200) distantly followed by Invasive lobular carcinoma (n=4). TNBCs with histological grade 2 have the highest frequency (112/200). The chi-square analysis showed that there was no significant association between histological type and tumour grade of TNBCs.

**Table IX:** Histological types and corresponding frequencies of the molecular subtypes of Triple negative breast carcinomas.

| **Variable** | **CK5/6 expression** |  | **AR expression** |
| --- | --- | --- | --- |
| **Histological type** | **Basal type** | **Non-basal type** | **Androgen receptor type** |
| Invasive carcinoma; NST | 53 | 134 | 36 |
| Mucinous carcinoma | 2 | 1 | 0 |
| Medullary carcinoma | 0 | 3 | 0 |
| Invasive lobular carcinoma | 1 | 3 | 1 |
| Invasive papillary carcinoma | 0 | 2 | 1 |
| Metaplastic carcinoma | 1 | 0 | 0 |
| **Total** | **57** | **143** | **38** |

This table shows that the most frequent histological type seen among patients with triple negative breast carcinoma is Invasive carcinoma; NST followed by invasive lobular carcinoma regardless of the molecular subtype.

**Table X:** Tumour sizes and corresponding frequencies seen among patients with triple negative breast carcinoma.

| **Tumour size** | **Frequency** |
| --- | --- |
| pT1 | 33 |
| pT2 | 39 |
| pT3 | 24 |
| pT4 | 7 |
| Unknown | 97 |
| Total | 200 |

This table shows that tumour sizes of ≥2.0cm and < 5.0cm (pT2) are the most frequent among patients with TNBCs seen in this study.

**Table XI:** Frequencies of tumour sizes among patients with triple negative breast carcinoma and lympho-vascular permeation.

| **Tumour sizes** | **Frequency** |
| --- | --- |
| T1 | 7 |
| T2 | 8 |
| T3 | 9 |
| T4 | 1 |
| UNKNOWN | 8 |
| **TOTAL** | 33 |

This table shows that tumour sizes of ≥ 5.0cm but < 10.0cm are the most frequent among patients who have lympho-vascular permeation

**Table XII:** Table showing the frequencies of the different molecular subtypes of TNBCs among those with lympho-vascular permeation.

|  |  | Molecular subtypes |  |  |
| --- | --- | --- | --- | --- |
| **Tumour sizes** | **Variable** | **Basal type** | **Androgen receptor type** | **Mesenchymal type** |
|  | T1 | 1 | 3 | 3 |
|  | T2 | 2 | 1 | 5 |
|  | T3 | 1 | 3 | 5 |
|  | T4 | 1 | 0 | 0 |
|  | Unknown | 1 | 2 | 5 |
|  | **Total** | **6** | **9** | **18** |

This table shows that the mesenchymal subtype was most frequent among those with lymphovascular permeation and are more of pT3 (tumour ≥ 5.0cm but < 10.0cm).

## PHOTOMICROGRAPHS

**Figure 4:**
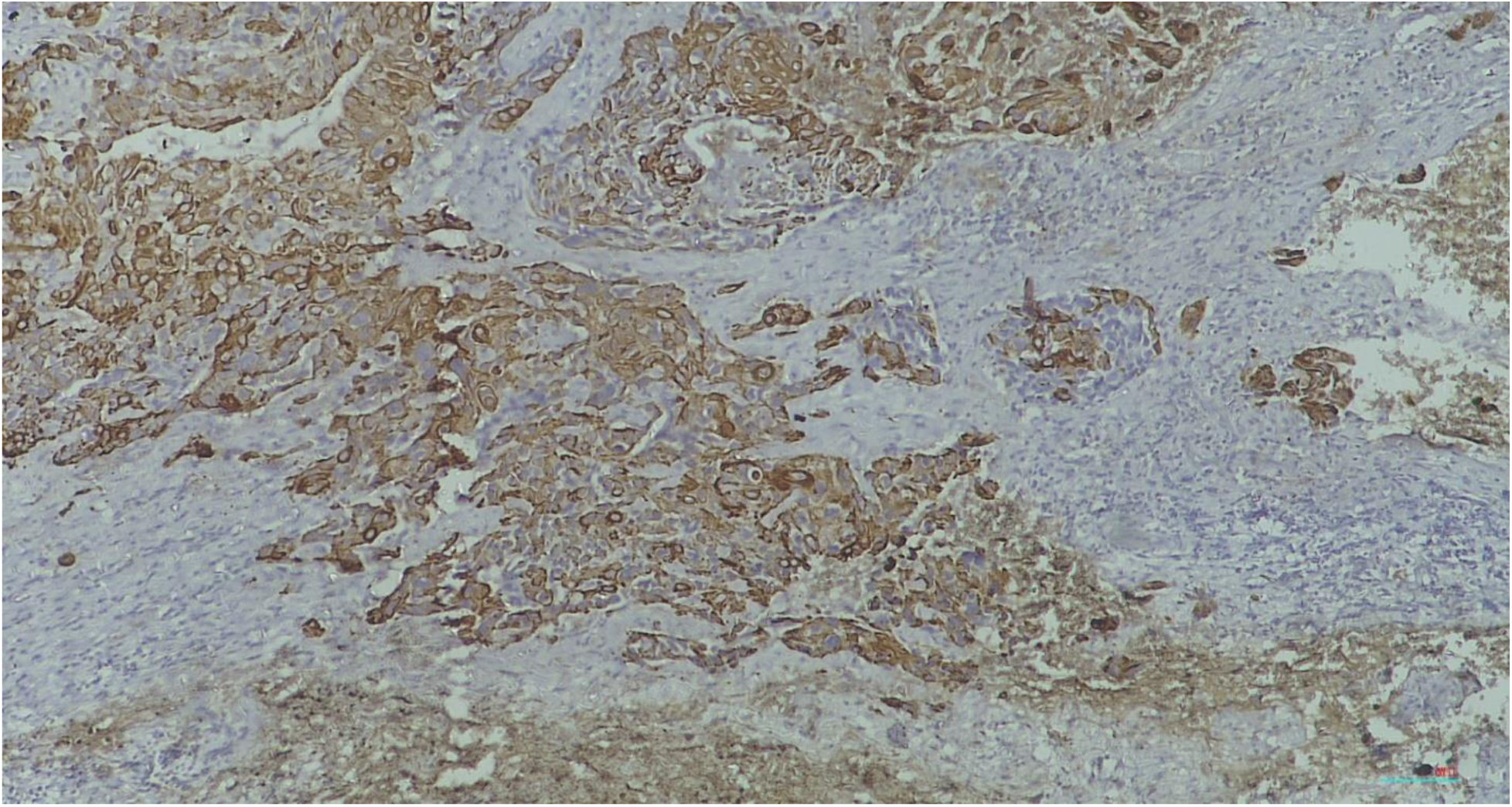
Photomicrograph (x100) of CK5/6 control slide using squamous cell carcinoma.

**Figure 5:**
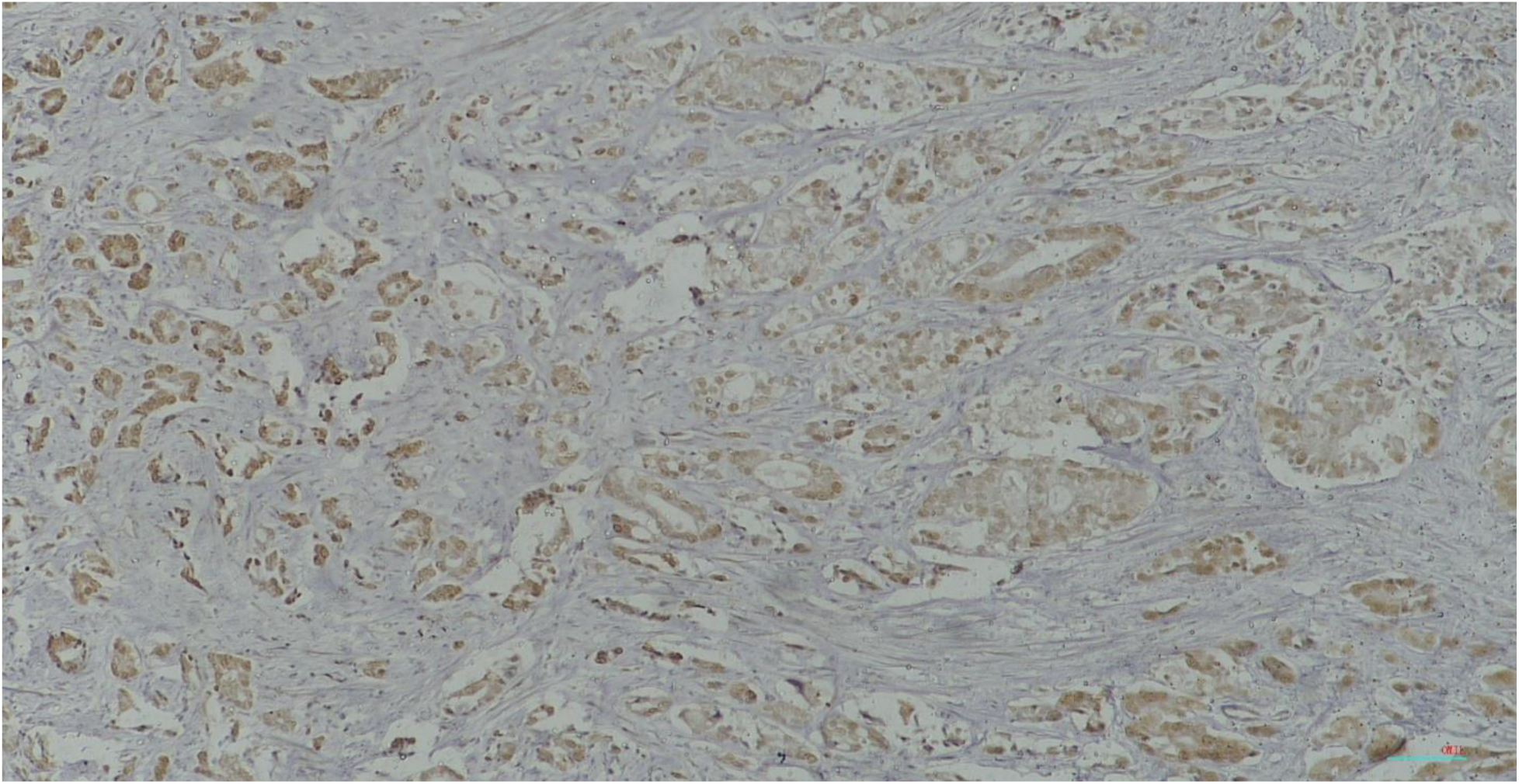
Photomicrograph(x100) of androgen receptor control slide using prostate carcinoma.

**Figure 6:**
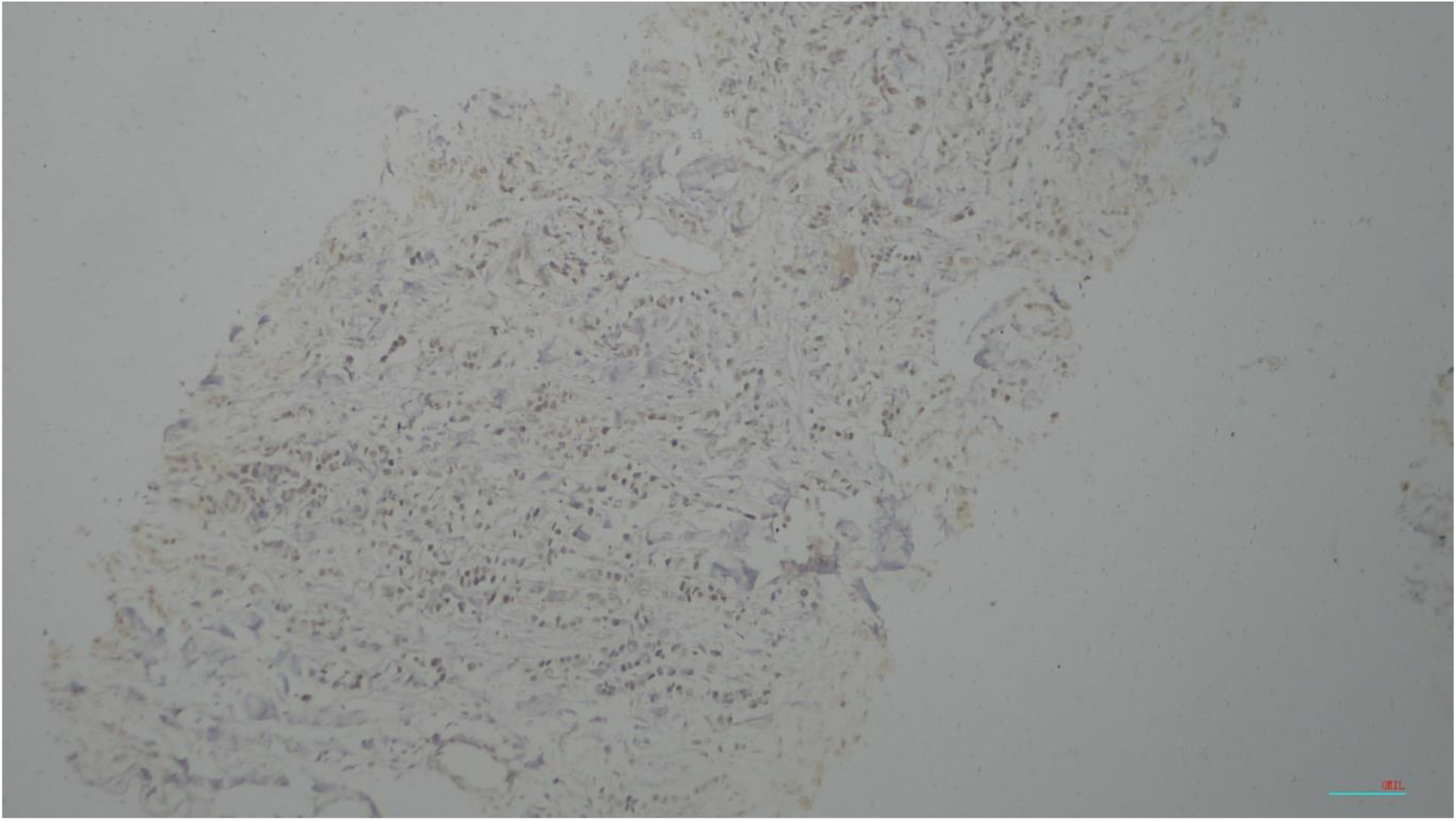
Photomicrograph (x100) LAR type of TNBC demonstrating androgen receptor positivity.

**Figure 7:**
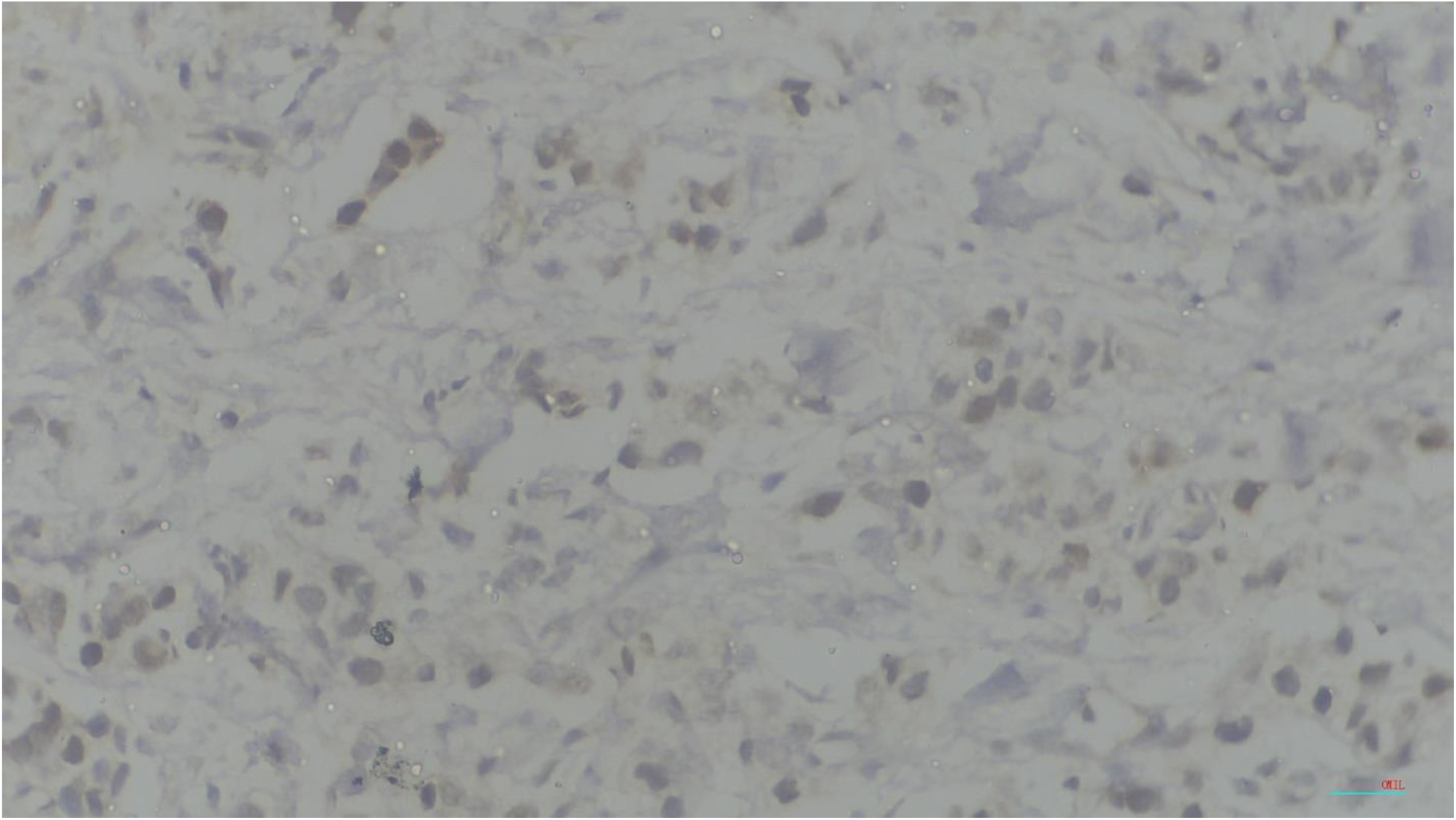
Photomicrograph (x400) LAR type of TNBC demonstrating androgen receptor positivity.

**Figure 8:**
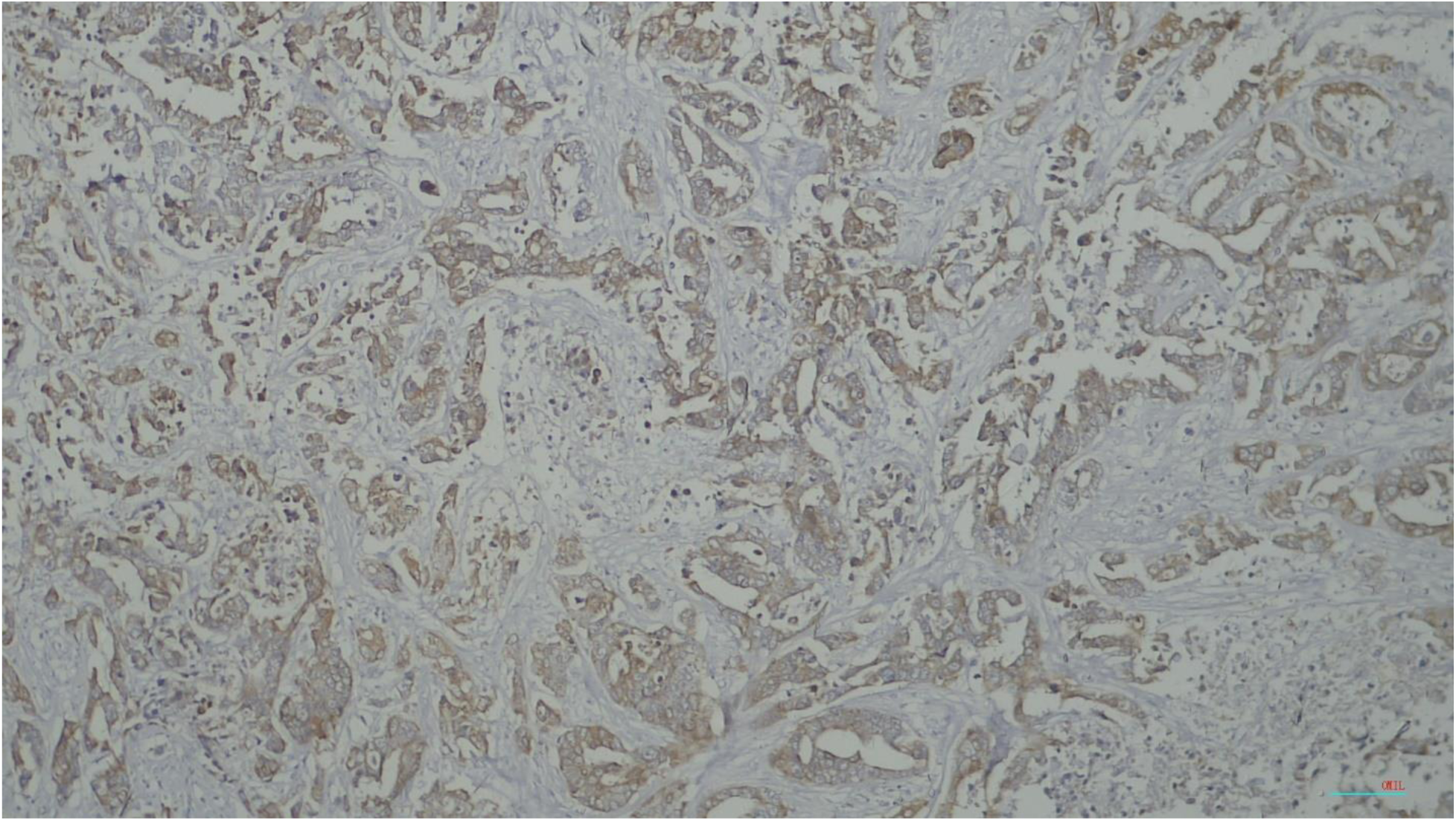
Photomicrograph (x100) basal type of TNBC demonstrating CK5/6 positivity.

**Figure 9:**
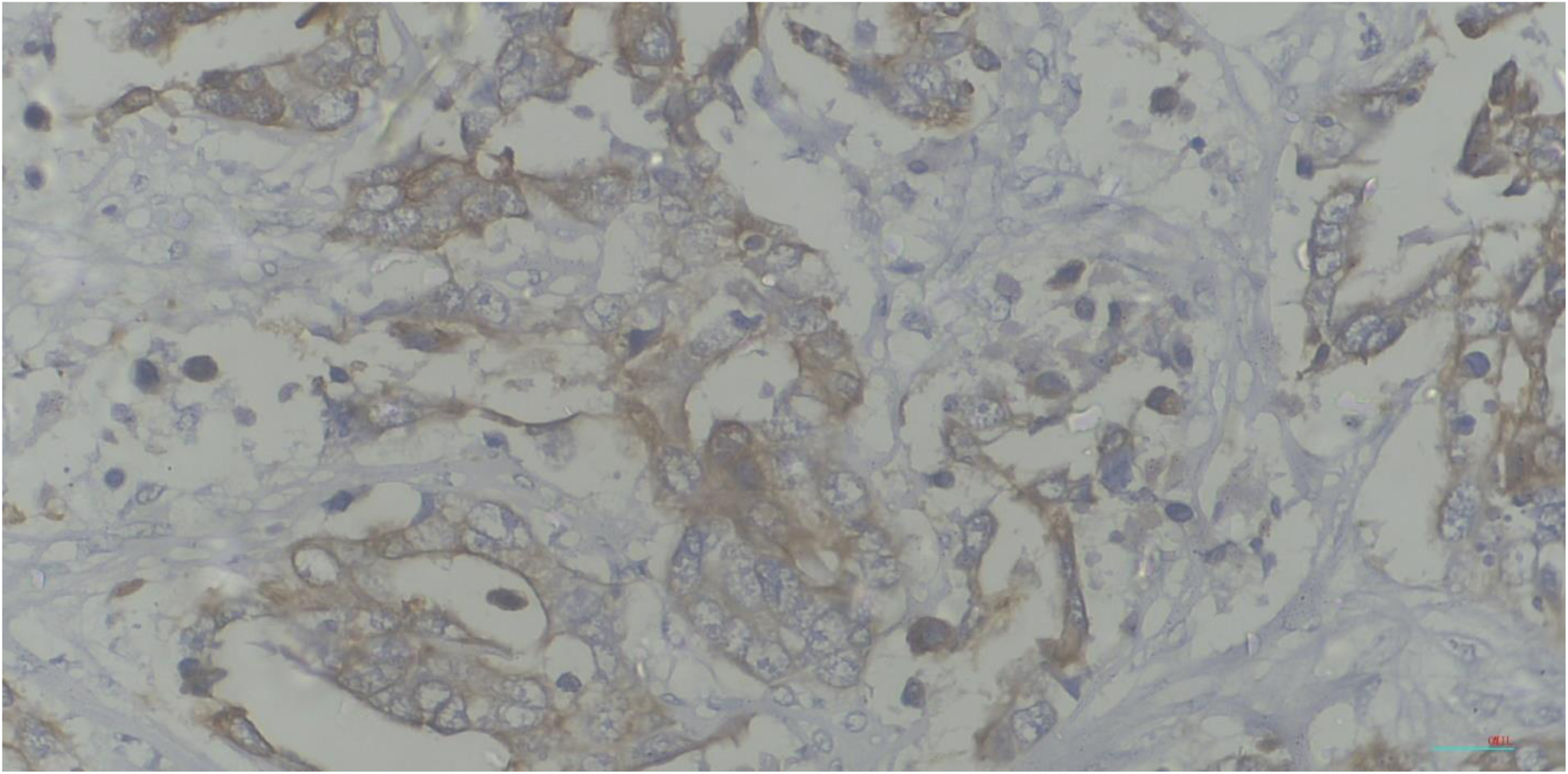
Photomicrograph (x400) basal type of TNBC demonstrating CK5/6 positivity.

**Figure 10:**
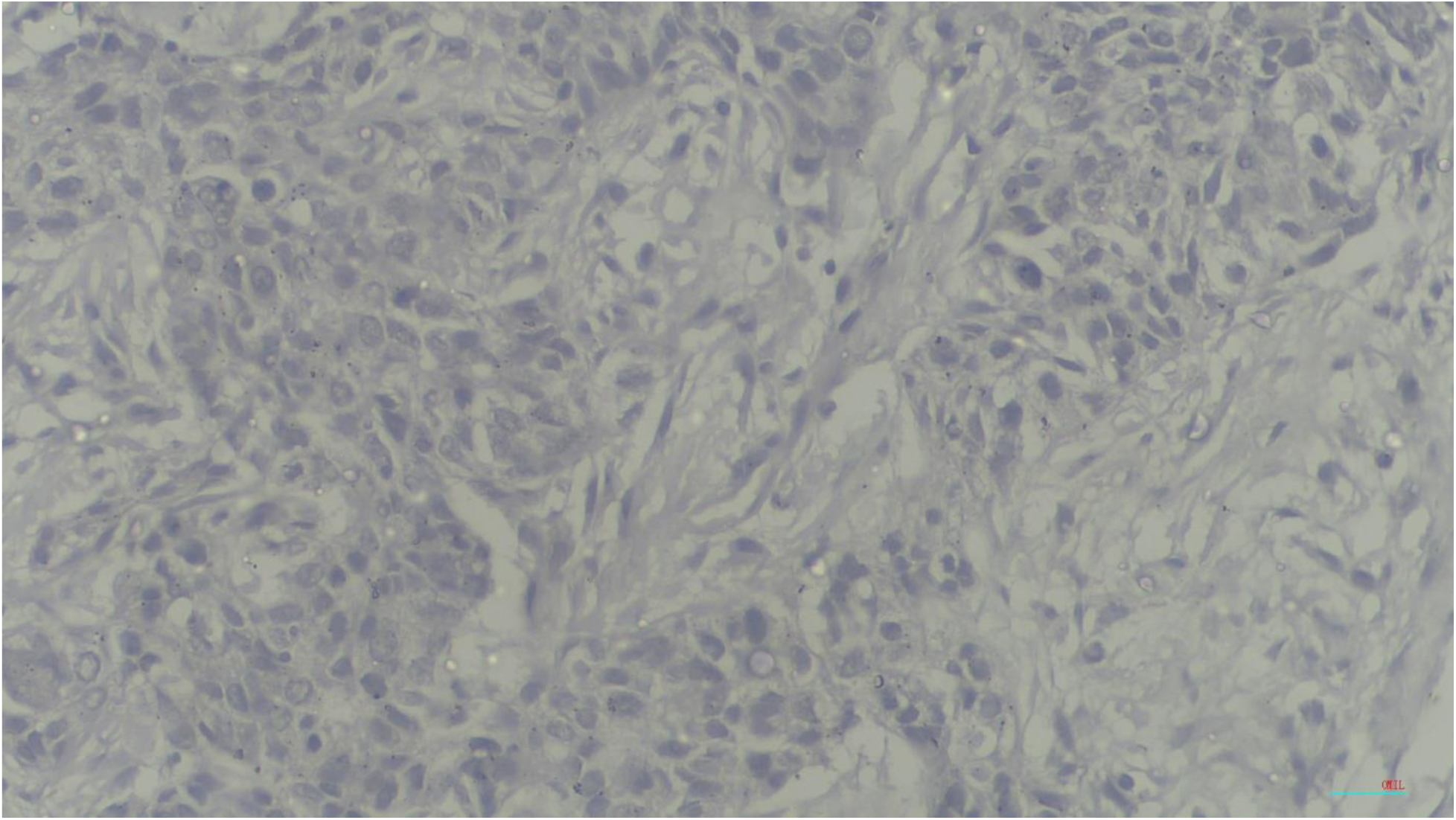
Photomicrograph (x400) non-basal TNBC demonstrating CK5/6 negativity.

**Figure 11:**
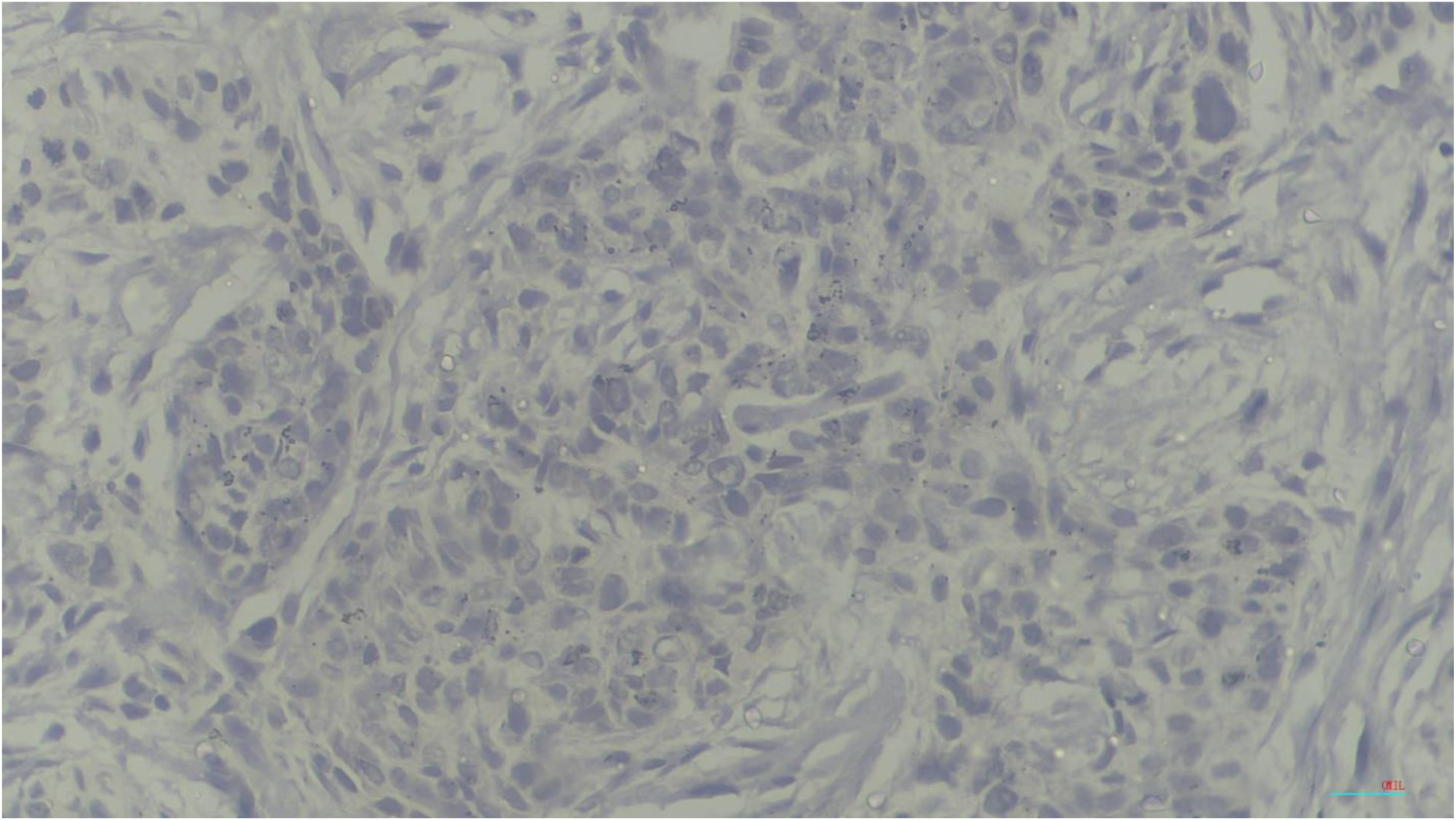
photomicrograph (x400) TNBC demonstrating androgen receptor negativity.

## CHAPTER FIVE

### 5.0 Discussion

Triple negative breast carcinomas in sub-Saharan Africa and in Nigeria has a high frequency and the frequency has been on a steady increase. Although it is a known fact that TNBCs have a poor prognosis, other factors have been observed through studies that increase the risk of recurrence and reduce the disease-free survival in patients. These factors include tumour size, lympho-vascular invasion, lymphocytic infiltrates, nodal involvement, visceral metastasis, age, Nottingham grade, expression of basal cytokeratins and androgen receptor^29,49^. Some of these factors are considered in this study.

### 5.1 The frequency of triple negative breast carcinomas

In this study, the relative frequency of TNBCs is 19.12 % which is far lower than that reported in other studies done in this environment (28.0 % to 65.0 %) ^10, 11^. This finding is similar to that of two Nigerian studies, Adebamowo et al and Zheng et al together with a study by Sayed et al^48, 49,50^ which reported TNBCs as 15.80 %, 11.7% and 20.2 % respectively. However, in slight contrast, studies in Asia, Europe and the Americas have shown slightly lower frequencies of TNBCs (12.18 %, 8.90 % and 12.50 % respectively) ^51–53^. These observed differences could be as a result of pre-analytical and analytical issues as of the time of those studies when immunohistochemistry was not routinely done in this country. In addition, population demographics in other continents reported more of post-menopausal age group in their studies and hence a higher proportion having breast carcinomas than the reproductive age group^19, 54^, in contrast to the findings in this environment where breast carcinoma is most commonly seen among the younger age group^38,55^. This may be explained by the higher frequency of *BRCA1* gene mutation in this region compared to that seen in developed countries ^17,56^. Also noted are variations in different studies conducted in Africa which although is within the same continent, epidemiological data on TNBCs is slightly different. This could be as a result of population-specific differences observed in some of these regions i.e. presence of mixed racial groups within those African countries like Anglo-Europeans or Americans residing within those countries and whose data would be considered as ones from Africa. Another influence on the frequency of TNBCs in this study though high, it is significantly lower than other previous reports which have higher frequencies in this region is the sample size. Only 62.0% of the total number of breast samples received during this study period were eligible for inclusion criteria which could subsequently also affect the skewing of samples regarded as TNBCs and non-TNBCs in this study. Wide variations in sample sizes and inclusion and exclusion criteria used in the different studies conducted in Nigeria is also noted as a possibility for the marked variations in frequencies of TNBCs.

### 5.2 Age distribution associated with triple negative breast carcinoma

The mean age of triple negative breast carcinomas in this study is 45.52 ±11.80 which is similar to an earlier study done in Maiduguri with a mean age of 45.4 ± 13.6SD years^34^ and a study in Asia^57^. This implies that majority of TNBCs diagnosed in this institution are within the age of 40-49 years as has been demonstrated. It was found that the 31-49 age group have the highest frequency of TNBCs which is also similar to the age group commonly affected by invasive breast carcinomas in this region^58,59^. In contrast, Bauer et al^52^ reported a mean age of 54 years and the highest frequency among the 50-59 age group. In addition, Pistelli et al^60^ in Europe and Naher et al^61^ in Australia have both reported a higher frequency of TNBCs in those > 50 years (post-menopausal), with a high grade (Grade 3) and high proliferative index though, many of the patients were diagnosed at the early stage of the carcinoma (Stage 1). This variance in the findings could be due to population-specific differences in tumour biology and environmental exposures that could cause carcinogenesis^13^. A study (Abdulrahman & Rahman, 2012) has shown that women with African ancestry have higher levels of oestradiol and lower levels of the sex hormone binding globulin. This therefore translates into higher risks and subsequently frequencies of breast cancers in Africa. Moreover, the population demographic in Nigeria is comparatively young^38^. This culminates in the skewing of data among the younger age groups in contrast to developed countries where life expectancies are higher.

### 5.3 The molecular subtypes of Triple negative breast carcinomas

In this study, frequencies of the basal and non-basal types are 28.5 % and 71.5 % respectively. This is at variance with the findings of Hubalek et al^17^ in Austria with higher frequencies of the basal than the non-basal type. The report in this study is however similar to that in Ibadan with a higher frequency of the non-basal type^38^. The non-basal type comprising of the androgen receptor and mesenchymal types constitute 19.0 % and 52.50 % respectively. This is also similar to a study by Fioretti et al^41^ which classifies AR predominantly in the non-basal group. In a multi-centre study involving six countries representing Africa (Nigeria), Asia (India), Europe (UK, Norway and Ireland) and the Americas (United States), the frequencies of androgen positive breast carcinomas were as follows; 8.30 %, 22.70 %, 54.90 %, 32.70 %, 24.70 % and 24.80 % respectively. The lowest frequency of the androgen receptor positive breast tumours is seen in the Nigerian centre and highest in the UK centre^43^. The frequency of AR positive breast carcinomas in this current study and the previous study from the Nigerian centres are both lower than that seen in the other regions. The lower frequencies seen in this continent could be attributed to demographic differences between Nigeria and developed countries where breast carcinoma cases are seen commonly in the older age group^11,38^. This is however in sharp contrast to that seen in this region where the occurrence in the younger age group (< 50 years – constituting 69.3 % in this study) is more frequent and closely associated with a higher stage and proliferative index^13^. Variations in reports could be because it has been established that AR positivity is more common in older women,^62^ and AR is only expressed in 30% of *BRCA1*-related TNBCs. The low frequency of AR positive tumours in this study is in support of other studies in this region that have reported a high frequency of *BRCA1* and *BRCA2* (though not as frequent) mutations in triple negative breast cancers^63^. In addition, AR expression has been reported to be higher in breast carcinomas of lower histological grades and lower stages as seen in developed countries than in carcinomas with higher histological grades and stages commonly seen in this region^43,64,65^. Furthermore, it has been found that physical characteristics of a population also have a strong association with TNBCs being more of the basal or non-basal type. A study in North-America among African-American women^24^ has shown a strong association between waist-hip ratio (which is a strong indicator of obesity and one of its indicators) and basal-like expression of TNBCs. The risk of basal-like expression was also increased in pre-menopausal women in the same study. This is however in contrast to this current study which has shown that despite the higher frequency of pre-menopausal women being affected, the non-basal type is the most frequent subtype in this study. Moreover, the same study has shown that the morphology of obesity in those with African ancestry is quite different from Europeans which has also been attributed to causing the expression of more aggressive forms of TNBCs^24^. In addition, a contrast to this study, a double cohort study^66^ has shown a positive correlation between *p53* mutation which is frequent in Nigerian premenopausal women and the expression of the basal phenotype. Though many studies regarding *p53* are yet to be conducted in Nigeria in association with TNBC, the non-basal type is most frequent in this study. Another important finding in this study is that there is no overlap between the expression of the androgen and basal cytokeratin expression. This is however in contrast to a study in the Americas^67^ which showed some overlap in 31.8% of the samples. An asian study^68^ has also shown that there is an inverse relationship between androgen receptor expression and basal markers. This was explained by high frequencies of *Rb* gene loss in this region compared to Asia leading to higher histologic grades and reduced AR expression as seen in this study. Another study in Asia^62^ showed dual positivity for androgen receptor and CK5/6 and classified those group of tumours as intermediate risk between the androgen receptor type which has the best prognosis and the basal type which has the worst prognosis. More studies however need to be conducted in this region to ascertain the relationship between *Rb* loss, androgen receptor and CK5/6 expression.

### 5.4 The frequencies of the histological types seen among patients with invasive breast carcinomas generally

In this study, Invasive carcinoma; Non-Specific Type (NST) is the most frequent histological type with a relative frequency of 95.8 % (n=1002/1046). This finding is similar to studies from Nigeria and other parts of the world^38,69–73^. However, the other special types of breast carcinomas seen vary in the different studies. The second most frequent in this study is Mucinous carcinoma. This is similar to what was reported by Jimoh et al in Ibadan^74^ and Rambau et al in Tanzania^58^. On the contrary, a study by Moudiongui et al in Congo Sub-Saharan Africa reported invasive lobular carcinoma as the second most seen histological type^75^. Other special types are seen in other studies that is not seen in this study^73,76^. Overall, the expression of other histological types apart from Invasive carcinoma; NST could be largely dependent on the population-specific differences or epigenetic modifications peculiar to other regions^77^. Furthermore, a study in Europe^78^ has also shown that majority of triple negative breast cancers are of invasive carcinoma; NST histological type followed by other rare types which are frequently triple negative inclusive of medullary and metaplastic carcinomas as seen in this study.

### 5.5 Frequencies of the histological grades seen in patients with invasive breast carcinomas

The most frequent histological grade is Grade 2. This is followed by Grades 3 and 1. This is similar to some studies done in Nigeria^65,68^ but in contrast to studies done by Hadgu et al^79^ in Ethiopia and Al-thoubaity^80^ in Saudi-Arabia in which Grades 3 and 1 tumours were the most common respectively. Variations in observations could be because a report has shown that worldwide, 60.0 % of tumours are readily classified as Grade 2^81^.Other contributing factors could include late diagnosis resulting from poor access to health care, low educational status affecting health-seeking behaviour, demographic differences in protein expression by region responsible for tumour aggressiveness^66,77,78^ and a strong association between TNBCs and *BRCA1* mutation^17,78^. A study has also shown an association between a high prevalence of Human Papilloma virus, HIV and hepatitis viruses and TNBCs though the exact relationship has not been ascertained. This has been proposed as a possible cause for tumour aggressiveness^77^.

### 5.6 Correlation between histological types and grades of patients with triple negative breast carcinomas

In this study, there is no correlation between the histological types and grades of triple negative breast carcinomas. Even though there is no correlation, a trend has been observed. Individual studies in Nigeria and Kenya have reported TNBCs to be more likely of Grade 3^82,83^. Reports that show correlation reported a higher frequency of grade 2 carcinomas than Grade 3 as seen in this study^33,84^. Some other studies in the Americas, Africa and Asia show varying correlation between histological type and grade^23,77,78^. The correlation seen in the African studies could be because it has been reported that TNBCs in those of black race are usually of a higher histological grade and advanced stage^33,77^. In addition, differences in protein expression associated with a higher histological grade among those of the black race compared to that of developed countries has been reported^82^. Moreover, no significant association was established (table VIII) between histological types and grades of TNBCs. This may be because globally, invasive carcinoma; NST is the most common histological type ^85^ and the diagnosis of TNBC confers a poor prognosis regardless of the histological type ^86^.

### 5.7 Other biographic data

In this study, though the overall expression of androgen receptor is not high, it is observed that the highest expression was found within the same age group with the highest frequency of TNBC, the 30-49 age group. This is in contrast to a Latin-American study where the mean age for AR positive TNBCs was the 46-62 age group^87^. This could be as result of the older age group in this population being more at risk of breast carcinoma than the younger age group.

In this study, there was some association observed between the histological types and the molecular subtypes of triple negative breast cancer (Table IX). In this study, 68.5% of the invasive carcinoma; NST type falls within the non-basal group (n=137/200), 20.5% were of the basal subtype (n=41/200), the androgen receptor subtype constitutes 18.0% (n=36/200). This is followed by invasive lobular carcinoma with 1.0% of the samples being of the basal and non-basal types respectively. The only metaplastic carcinoma in this study is of the basal type. 0.5% of the invasive papillary carcinoma type was of the androgen receptor subtype and 1.0% was negative for both the androgen receptor and CK5/6. Data regarding association between the different histological types and molecular subtypes of TNBCs are not readily available. However, a close study to this in Nigeria is that done by Liman et al^88^ who only established the different histological types found within the luminal androgen receptor subtype.

There were variations in tumour sizes seen in this study. 73.3% of the samples were of pT1 and pT2. This is in contrast to an African study and a Nigerian study ^85,89^ where 70.1% and 87.4% of the tumours were of pT2 and pT3 respectively. This finding also differs from that seen in an Asian study^90^ where 38.0% of the samples had tumour size less than 5cm and only 33.0% had tumour sizes greater than 5cm. However, in similarity to this study, a study in the Americas has shown a higher frequency of pT1 and pT2 tumours^91^. This could be as a result of the finding that despite the high frequency of TNBC in the Asian study (43.0%), majority of the samples (90.0%) in that study were intraductal carcinoma. Also, the high frequency of T1 and T2 tumour sizes in this study could be as a result of the high level of education among a large number of patients living in the city and also readily have access to the hospital and also influencing their health-seeking behaviour. Moreover, lympho-vascular permeation was assessed in 16.5% of the samples in this study which is also in contrast to the afore-mentioned African study in which 48.6% of the samples were positive for lympho-vascular permeation. This result stems from the finding that not all the tumours in this study could be assessed for lympho-vascular permeation coupled with incomplete data. Studies have shown a direct relationship between tumour size and increased risk of lympho-vascular permeation. In this study, the samples with lympho-vascular permeation had tumour sizes T3 and T2 at n=9/33 and n=8/33 respectively. This is followed by tumour size of T1. The T1 tumours had equal numbers of AR positive and mesenchymal subtypes (9.1% respectively), T2 tumours had a higher frequency of the mesenchymal type (15.2%) and basal type (6.1%) than the AR positive type (3.0%). Tumours with pT3 sizes had a higher frequency of the mesenchymal subtype (15.2%) and androgen receptor subtype (9.1%) than the basal types (3.0%). There was only one sample with T4 and of basal subtype in this study. This shows that pT2 and pT3 tumour sizes are more likely to have lympho-vascular permeation and higher chances of being of the mesenchymal and androgen receptor subtype. AR expression has been associated smaller tumour sizes but the contrast is found in this study^92^.

In this study, the invasive carcinoma; NST histological type was found in all the age groups represented in this study. The second most frequent histological type; mucinous carcinoma was found in the 30-39 and 60-69 age group. Invasive lobular carcinoma was seen in the 30-59 age group and invasive papillary carcinoma was found in the 40-49 and ≥ 70 years age groups. Medullary carcinomas was seen in the 30-49 age group. Some variations were seen between this study and an Italian study^93^. In contrast to this study, the various histological subtypes were most frequently seen among the post-menopausal age group. This is because breast carcinomas and TNBCs are most frequently diagnosed in the older age group than in this region.

### 5.1. Conclusion

This study has shown that triple negative breast carcinomas diagnosed in the Department of Histopathology National Hospital, Abuja constituted 19.12 % of all invasive breast carcinomas and were more commonly seen within the pre-menopausal age group (30-49 years). Patients with tumour sizes of ≥ 2.0cm but < 5.0cm (pT2) were most frequent among the TNBCs and tumour sizes of ≥ 5.0cm but < 10.0cm (pT3) were most frequent in those with lympho-vascular permeation within the study samples in the hospital. Among the samples that had data on tumour sizes in association with lympho-vascular permeation, the mesenchymal subtype was the most frequent followed by the androgen receptor subtype. The androgen receptor subtype accounted for 19.0 % of TNBCs. This work has shown that patients with androgen receptor subtype of TNBC will benefit from anti-androgen therapies as is the practice in centres outside Nigeria. It has also shown that patients can benefit from screening programs that will detect breast tumours early regardless of molecular type. Furthermore, this study has shown that the most frequent histological type is Invasive carcinoma; Non-specific type (NST) regardless of the molecular subtype and presence or absence of lympho-vascular permeation. This study has also shown that the most frequent histological grade seen among patients with TNBCs is Grade 2.

### 5.2. Recommendations

1. All TNBCs should be tested for androgen receptor positivity; this is for improved management outcome of patients.
2. Pre-analytical issues should be properly managed. This is to minimize errors to a large extent and provide room for accurate diagnosis in immunohistochemical tests.

## Data Availability

All data produced in the present work are contained in the manuscript.

## Acknowledgement

I thank God Almighty for his guidance, wisdom and mercies everyday of my life. I would like to thank all my teachers; both immediate and past ones who have helped shape me into the person I am today. Lastly, I cannot forget the support of my family and friends who stood by me through thick and thin with love, care and understanding. God bless you all.

## LIST OF ABBREVIATIONS

TNBCs: Triple negative breast carcinoma
C/K: Cytokeratin
HER2: Human Epidermal Growth Factor 2
WHO: World Health Organization
SPSS: Statistical Package for Social Sciences
EGFR2: Epidermal Growth Factor Receptor 2
CAP: College of American Pathologists
ASCO: American Society of Clinical Oncology
CEP 17: Chromosome Enumeration Probe 17
ER: Oestrogen receptor
PR: Progesterone receptor
SEER: Surveillance, Epidemiology and End Results
*BRCA*: Breast Cancer gene
AJCC: American Joint Committee on Cancer
EU: European Union
RCPath: Royal College of Pathologists
UK: United Kingdom
NOS: Not Otherwise Specified
NST: No Special Type
*Rb*: Retinoblastoma gene
*P53*: Tumour protein p53
*Ki67*: Marker of proliferation Ki-67
*TOP2A*: Topoisomerase (DNA) II alpha
*BRCA2*: Breast cancer gene 2
*BRCA1*: Breast cancer gene 1
BL-1: Basal-like 1
BL-2: Basal-like 2
IM: Immune-modulatory
M / MES: Mesenchymal
MSL: Mesenchymal stem-like
LAR: Luminal androgen receptor
BLIS: Basal-like immune-suppressed
BLIA: Basal-like immune-activated
*ATR/BRCA*: ataxia-telangiectasia-Rad3-related/breast cancer genes
PDGF: Platelet -derived growth factor
ERK1/2: Extracellular signal-regulated protein kinase
ATP: Adenosine triphosphate
*ERB-B2*: Erb-B2 Receptor Tyrosine Kinase 2 gene
DHT: Dihydrotestosterone
BAFs: Breast adipose tissue fibroblasts
FISH: Flourescent in-situ hybridization

## APPENDIX I

**TABLE I.** : Potential Biomarkers Underlying Nigerian TNBC^13^.

| **Genetic variant** | **Detection technique** | **Biomarker description** | **TNBC risk Association** | **Potential therapeutic agents** |
| --- | --- | --- | --- | --- |
| *Ancestry informative markers* | Genome-wide admixture mapping | SNPs associated with European or African ancestry | ER/PR-positive and early stage breast cancer associated with European AIMs |  |
| *TERT rs10069690 SNP in 5p15* | Genome-wide association study | Low-penetrance genetic variant; higher frequency of the allele in women of African ancestry | Strongly associated with ER-negative/PR-negative BC and TNBC |  |
| *BRCA1* mutation | Whole-genome sequence analysis | Tumour suppressor gene involved in DNA damage repair; associated with poor prognosis in BC and increased sensitivity to DNA damage agents | Associated with 50% increased risk for TNBC; prevalence is only approx. 1.4% in African-americans but approx. 7% among Nigerian patients with BC. | Platinum-based DNA damage agents (e.g, cisplatin, carboplatin), PARP inhibitors |
| Mutant *p53* | IHC | Tumour suppressor gene involved in | Higher expression in Nigerian BC | p53 induction of massive apoptosis |
|  |  | DNA damage repair; associated with poor survival, metastasis and chemotherapy resistance in BC. | cases compared with grade-matched BC specimens from UK; positively correlated with ER/PR-negative BC, TNBC, basal-like phenotype, premenopausal status, younger age at diagnosis, larger tumour size. More frequent in Nigerian patients than in UK patients. | (PRIMA-1), WEE1 or CHK1 inhibition, and p53 mutant-specific inhibitors |
| *Aurora A* | IHC | Tumour suppressor and regulator of stem cell renewal | Overexpression associated with ER-negative/PR-negative status, TNBC, basal-like BC; expression correlated with down-regulation of E-cadherin and up-regulation of CK5/6 and Ki67 among | Aurora kinase inhibitors |
|  |  |  | Nigerian patients with BC1 |  |
| *Ku70/80* | IHC | NHEJ regulator; associated with higher grade, LVI and basal-like phenotype | Expression positively correlated with TNBC and basal-like phenotype, metastatic disease, and down-regulation of BRCA1 and p53; expression prognostic in Nigerian patients |  |
| *Ki67* | IHC | Cell proliferation biomarker; expressed higher in AA compared with EA patients with BC | Higher expression in Nigerian patients with BC compared with grade-matched UK patients with BC; expression positively correlated with TNBC status and/or basal-like tumour biology, down-regulation of E-cadherin, BRCA1 mutation and up-regulation of p53,CK5/6 and CK16. |  |
| *PARP1* | IHC | Repairs DNA damage(i.e. single strand DNA breaks) through poly-ADP ribosylation to promote cell survival | Expression positively correlated with ER-negative/PR-negative and TNBC status, basal-like BC, higher grade, CK5/6 and CK14 expression, p53, EGFR, Ki67, and PI3KCA expression; poor prognosis biomarker in Nigerian patients with BC | PARP1 inhibitors |
| *PIASy* | IHC | Inhibitor of STAT signalling pathway; E£ SUMO protein ligase; implicated in BRCA1 deficiency and TNBC status. | Expression positively correlated with high grade, expression of basal-like biomarkers, ER-negative/PR-negative status, TNBC, basal-like phenotype; |  |
|  |  |  | poor prognosis<br>biomarker in<br>Nigerian patients<br>with BC |  |
| *ALDH1* | IHC | Cancer stem- and<br>progenitor-cell<br>marker | Higher expression<br>in Ghanaian patients<br>with BC; expression<br>associated with<br>TNBC. | ALDH1 inhibitors |
| *EZH2* | IHC | Oncogene involved<br>in transcriptional<br>repression; strong<br>association with<br>ER/PR-negative and<br>metastatic BC and<br>poorer clinical<br>outcomes | Over-expression<br>positively correlated<br>with high tumour<br>grade and basal-like<br>phenotype;<br>cytoplasmic EZH2<br>expression<br>associated with<br>TNBC among<br>Ghanaian patients<br>with BC | EZH2 and HMT<br>inhibitors |

## APPENDIX II

**TABLE II:**
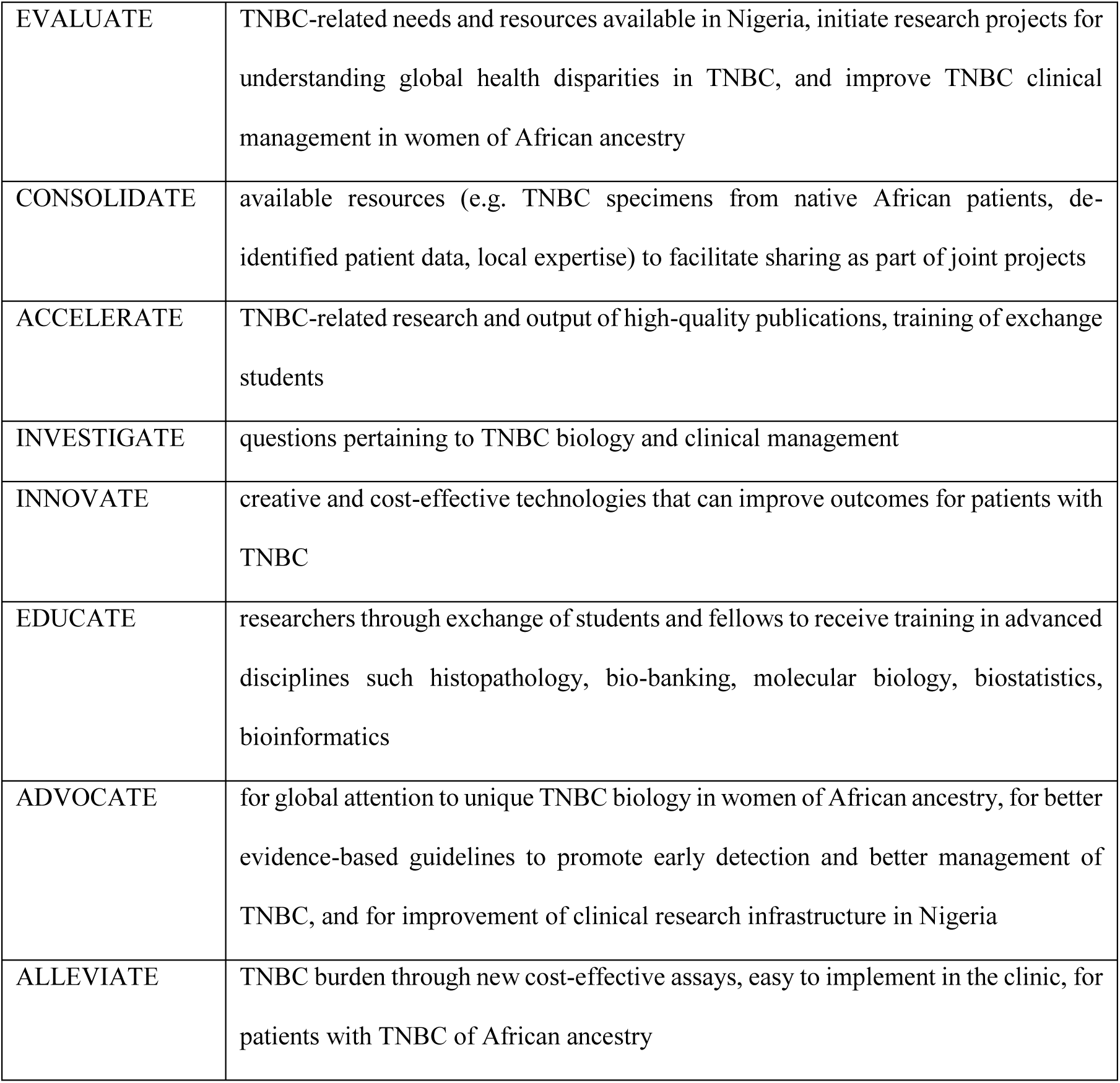
International Consortium for Advancing Research on Triple-Negative Breast Cancer (TNBC) strategic action plan for Nigeria^13^.

## APPENDIX III

**TABLE III:**
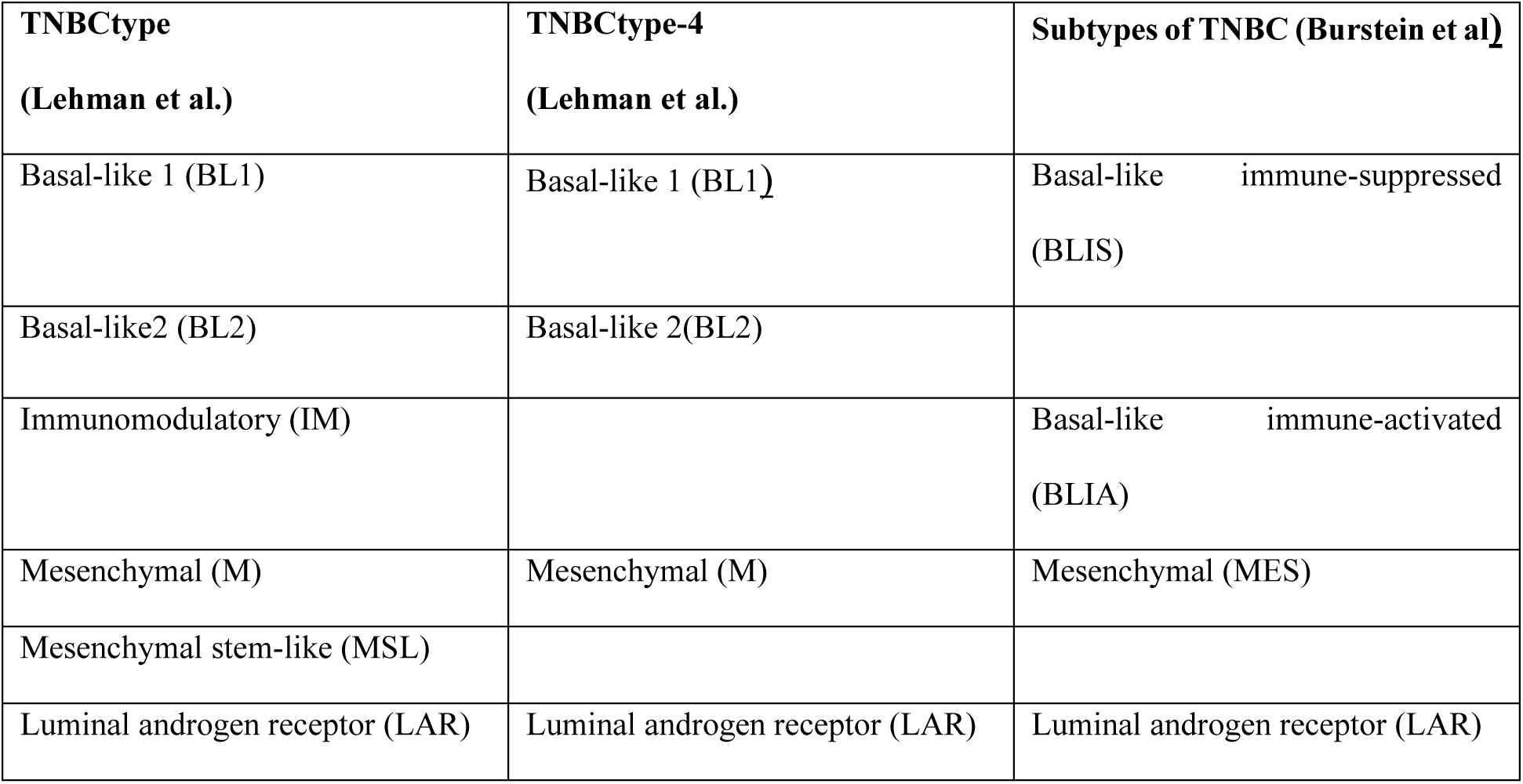
Subtypes of triple negative breast cancer (TNBC) based on analysis of gene expression profiles^21^.

## APPENDIX IV

**Fig. 1:**
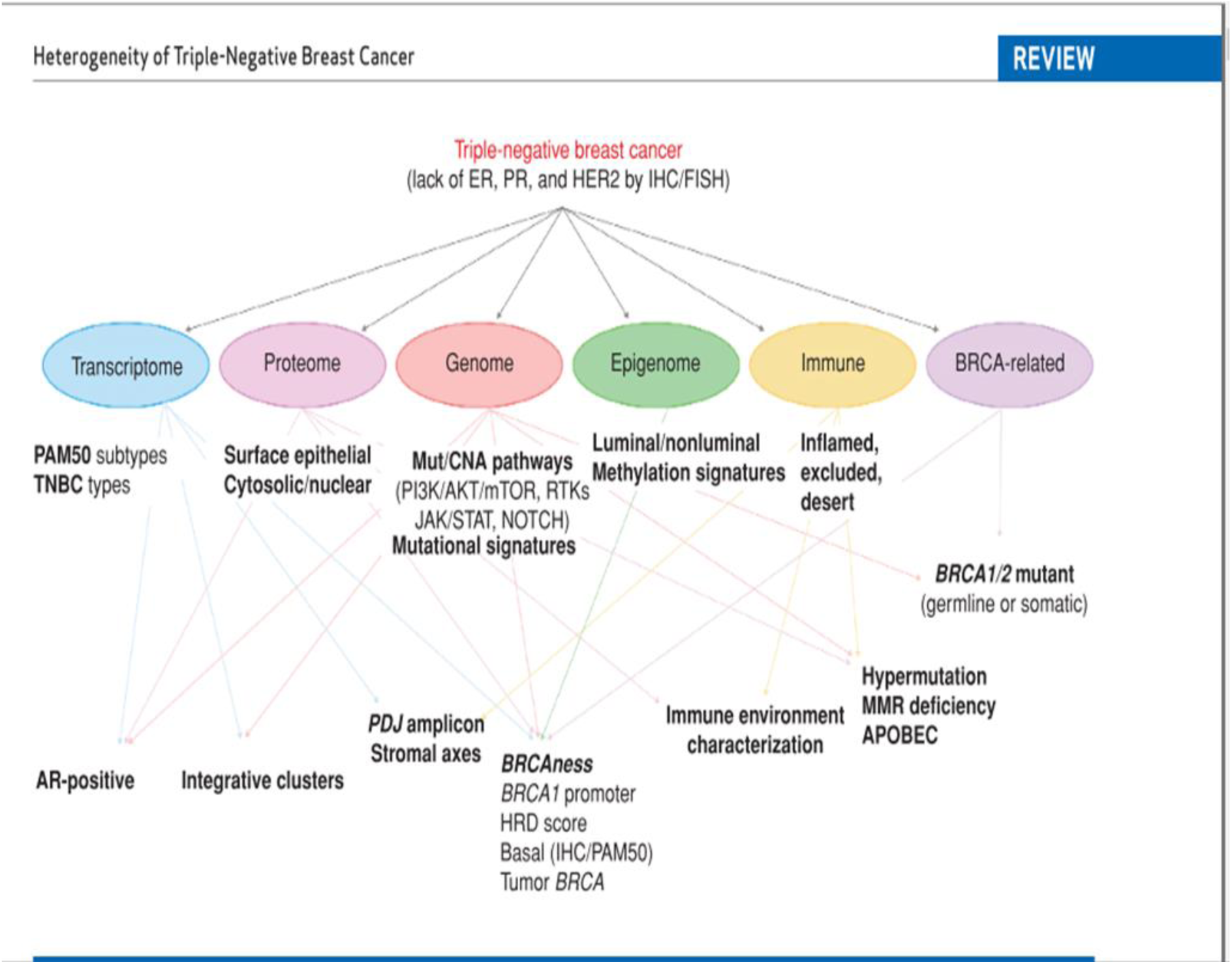
Parameters used in the sub-classification of TNBCs^36^.

## APPENDIX V

Tumour grading by the Nottingham upgrade of the Bloom-Richardson scoring system^29^

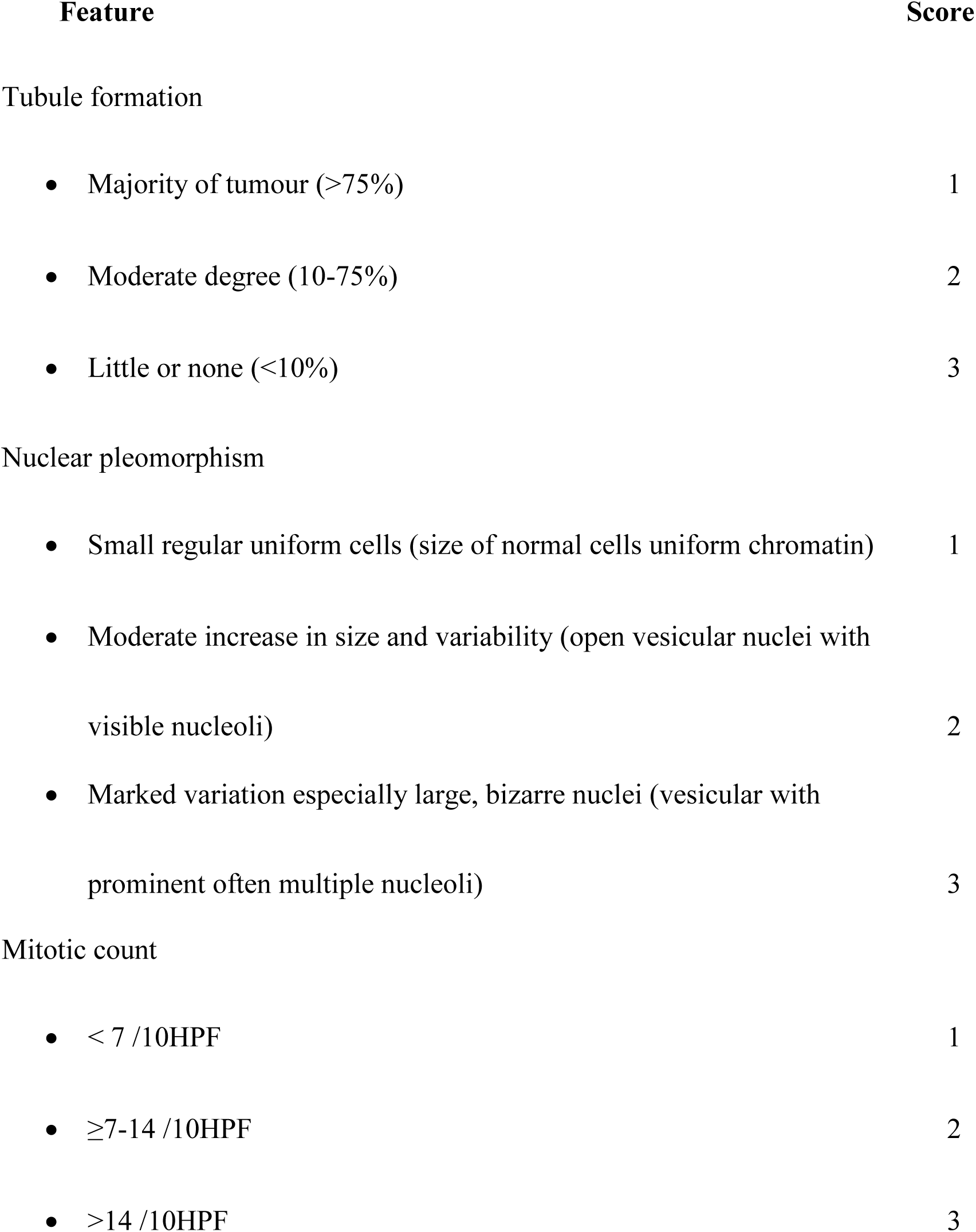
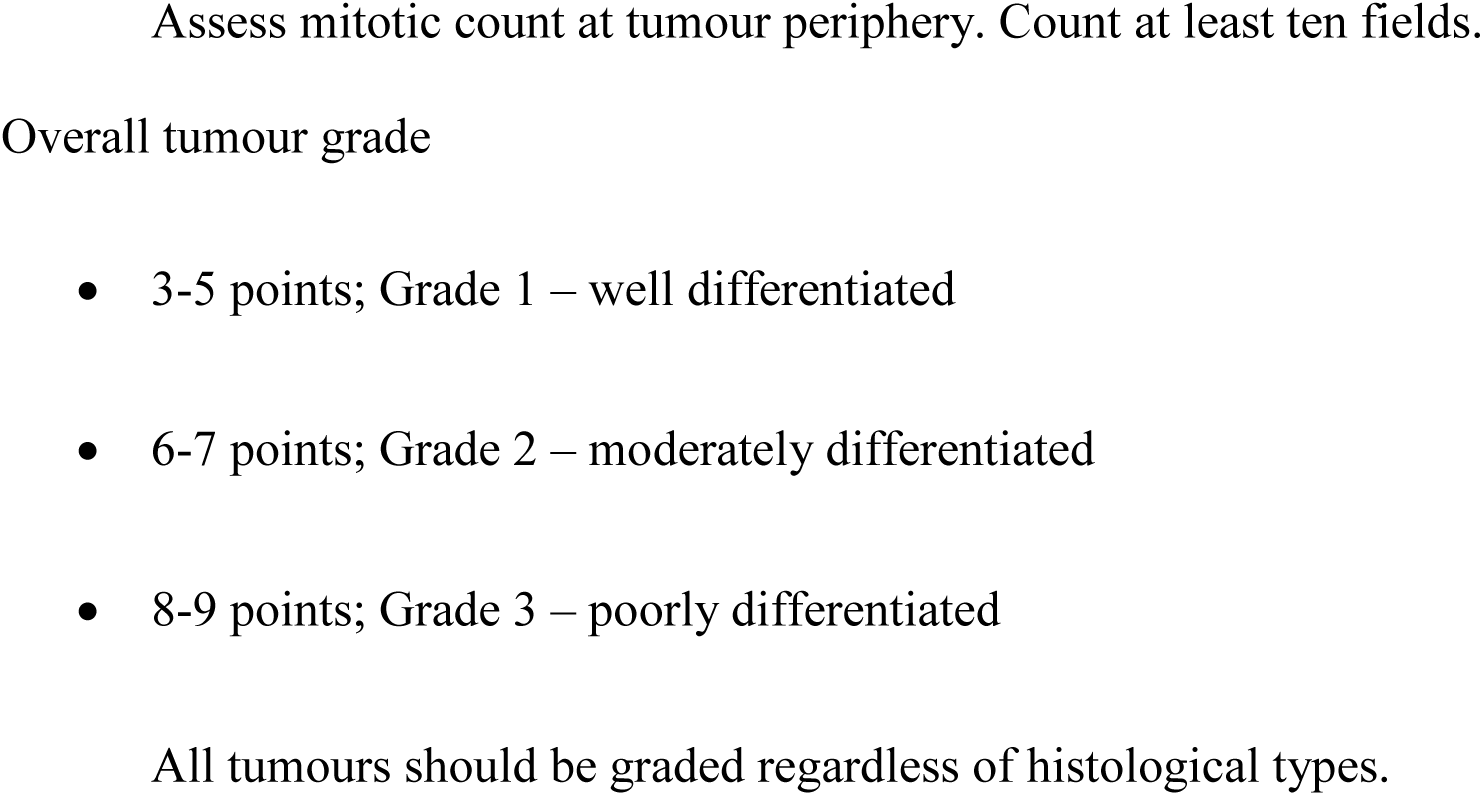

## APPENDIX VI

**TABLE IV.**
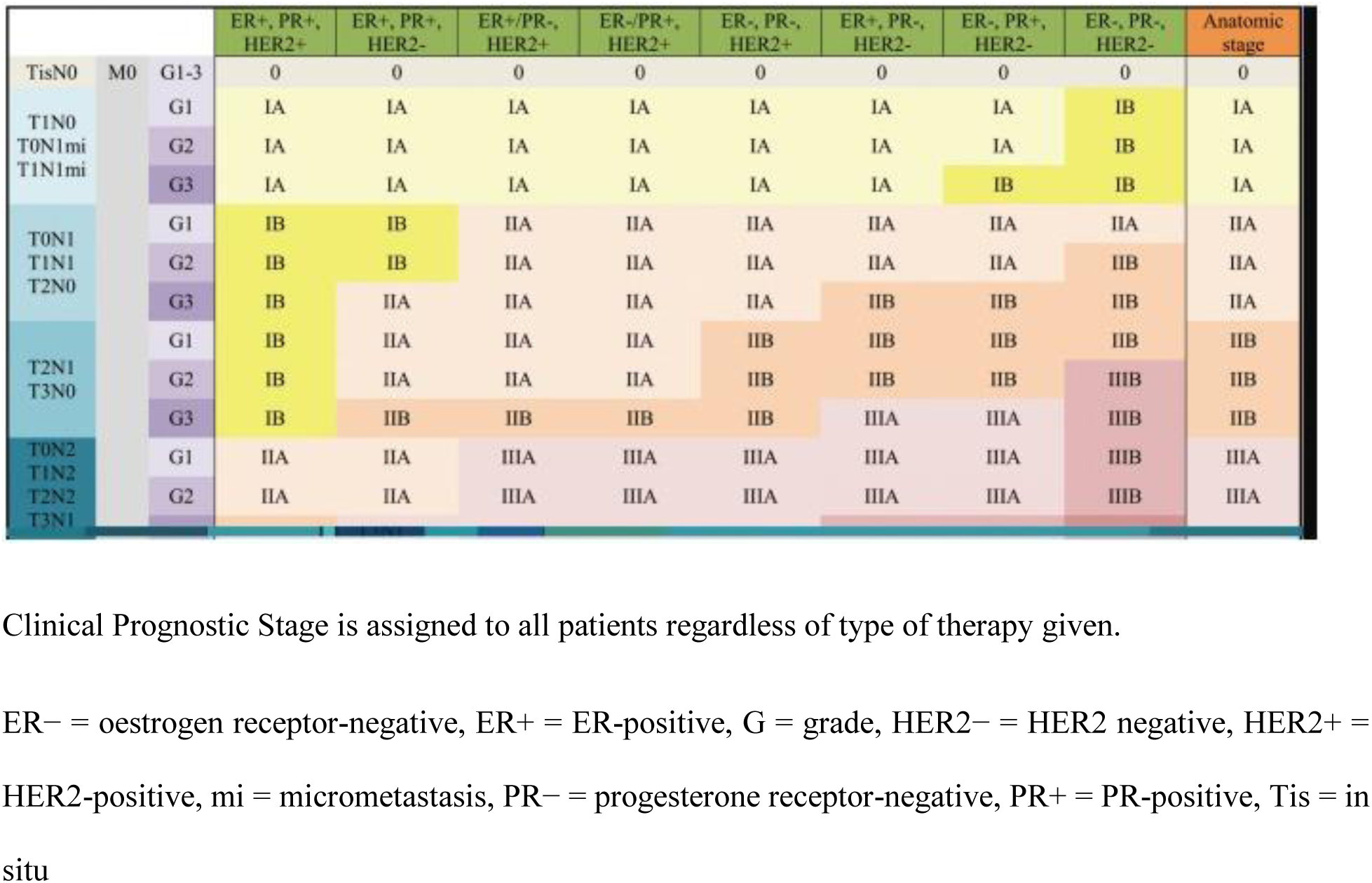
: Clinical Prognostic staging of breast cancer^46^.

## APPENDIX VII

**TABLE V.**
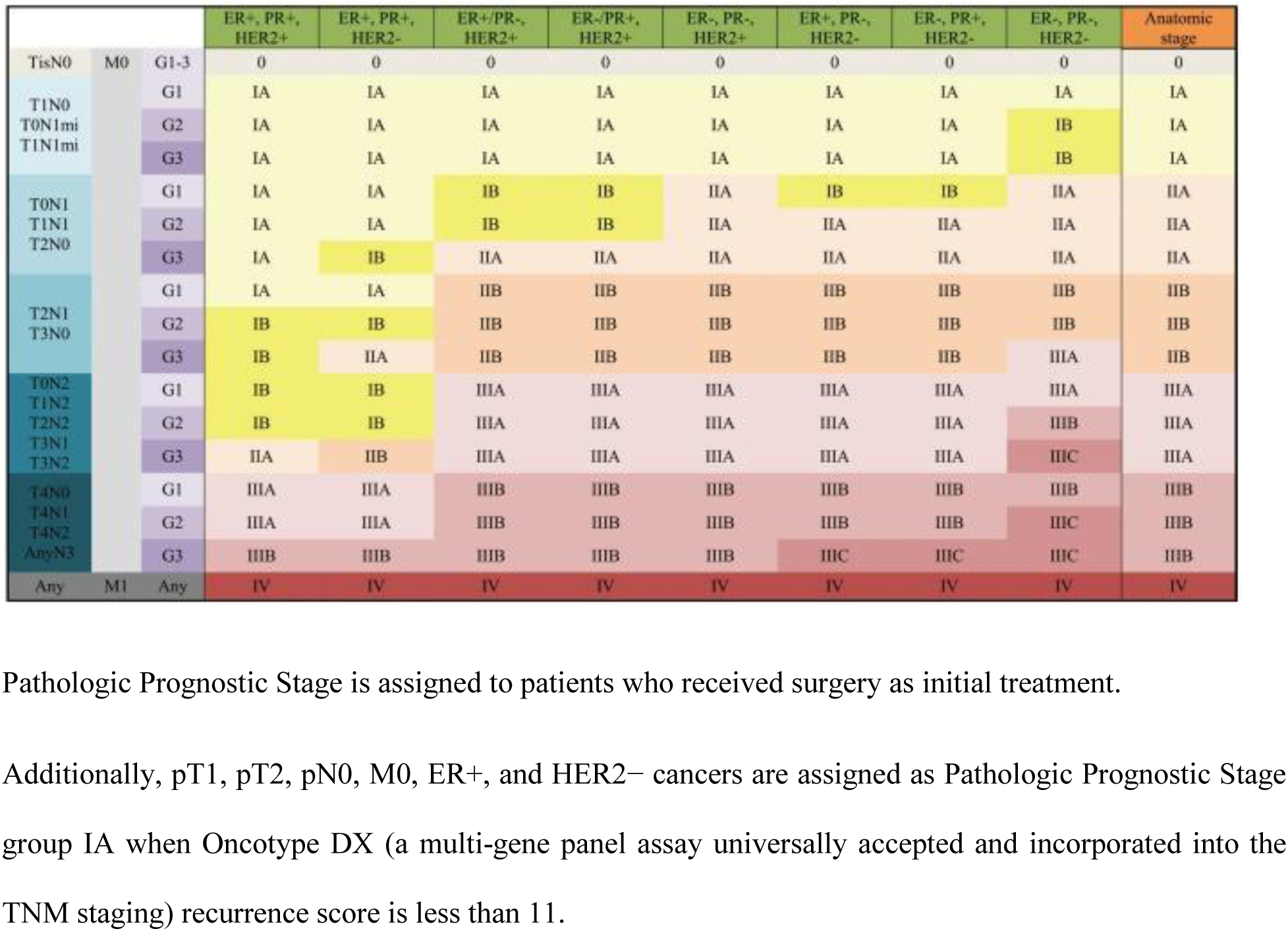
: Pathologic prognostic staging of breast cancer^46^.

## APPENDIX VIII

**TABLE VI:** WHO classification of breast tumours; 4^th^ Edition^47^.

| Type | Classification |
| --- | --- |
| Invasive carcinoma of no special type (NST) | 8500/3 |
| Pleomorphic carcinoma | 8522/3 |
| Carcinoma with osteoclast-like stromal giant cells | 8035/3 |
| Carcinoma with choriocarcinomatous features |  |
| Carcinoma with melanotic features |  |
| Invasive lobular carcinoma | 8520/3 |
| Classic lobular carcinoma |  |
| Solid lobular carcinoma |  |
| Alveolar lobular carcinoma |  |
| Pleomorphic lobular carcinoma |  |
| Tubulolobular carcinoma |  |
| Mixed lobular carcinoma |  |
| Tubular carcinoma | 8211/3 |
| Cribriform carcinoma | 8201/3 |
| Mucinous carcinoma | 8480/3 |
| Carcinoma with medullary features |  |
| Medullary carcinoma | 8510/3 |
| Atypical medullary carcinoma | 8513/3 |
| Invasive carcinoma NST with medullary features | 8500/3 |
| Carcinoma with apocrine differentiation |  |
| Carcinoma with signet-ring-cell differentiation |  |
| Invasive micropapillary carcinoma | 8507/3 |
| Metaplastic carcinoma of no special type | 8575/3 |
| Low-grade adenosquamous carcinoma | 8570/3 |
| Fibromatosis-like metaplastic carcinoma | 8572/3 |
| Squamous cell carcinoma | 8070/3 |
| Spindle cell carcinoma | 8032/3 |
| Metaplastic carcinoma with mesenchymal differentiation |  |
| Chondroid differentiation | 8571/3 |
| Osseous differentiation | 8571/3 |
| Other types of mesenchymal differentiation | 8575/3 |
| Mixed metaplastic carcinoma | 8575/3 |
| Myoepithelial carcinoma | 8982/3 |
| Epithelial-myoepithelial tumors |  |
| Adenomyoepithelioma with carcinoma | 8983/3 |
| Adenoid cystic carcinoma | 8200/3 |
| Rare types |  |
| Carcinoma with neuroendocrine features |  |
| Neuroendocrine tumor, well-differentiated | 8246/3 |
| Neuroendocrine carcinoma poorly differentiated (small cell carcinoma) | 8041/3 |
| Carcinoma with neuroendocrine differentiation | 8574/3 |
| Secretory carcinoma | 8502/3 |
| Invasive papillary carcinoma | 8503/3 |
| Acinic cell carcinoma | 8550/3 |
| Mucoepidermoid carcinoma | 8430/3 |
| Polymorphous carcinoma | 8525/3 |
| Oncocytic carcinoma | 8290/3 |
| Lipid-rich carcinoma | 8314/3 |
| Glycogen-rich clear cell carcinoma | 8315/3 |
| Sebaceous carcinoma | 8410/3 |

## APPENDIX IX

**TABLE VII:** Details of Antibodies used in this study.

| **Antibody** | **Source** | **Clone** | **Dilution** | **Control** | **Staining** | **Cut-off** |
| --- | --- | --- | --- | --- | --- | --- |
| Androgen<br>receptor | Biocare<br>medical<br>(mouse<br>monoclonal) | AR441 | 6.0mls,<br>prediluted (x<br>2) | Prostate<br>Carcinoma | Nuclear | $\geq 1.0\%$ |
| Cytokeratin<br>5/6 | Invitrogen<br>(mouse<br>monoclonal) | D5/16B4 | 1:100 | Squamous cell<br>carcinoma of the<br>skin | Membranous | $\geq 10.0\%$ |

## APPENDIX X

### Immunohistochemistry Protocol

Reagent preparation: Biocare medical (for androgen receptor), thermo-fisher antibodies (for cytokeratin 5/6) and Biogenex detection kit were used. All reagents were prepared according to the manufacturer’s instructions.

#### Deparaffinization and rehydration

**1.** Slides are incubated for 10 minutes at room temperature
**2.** Slides are subsequently rinsed in alcohol for 2 minutes and de-ionised water for 5 minutes

#### Heat-induced antigen retrieval (HIAR)

**1.** The deparaffinized slides are placed in plastic Coplin jar and heat retrieval done using the Biocare’s Borg decloaker.
**2.** The slides are allowed to cool gradually under running water for 2 to 5 minutes

#### Staining procedure

1. Peroxidases in tissues are blocked using Horse-radish Peroxidase block for 5 minutes at room temperature
2. Slides are washed in distilled water and then Tris-buffered saline (TBS)
3. Slides are incubated for 30 minutes at room temperature
4. The slides are washed off with TBS
5. The slides are incubated with the secondary antibody for 10 minutes at room temperature
6. The slides are washed off with TBS
7. For the androgen receptor, the slides are incubated for 20-30 minutes at room temperature with a tertiary polymer.
8. Slides are incubated for 5 minutes at room temperature with Diaminobenzidine (DAB)
9. The slides are washed off with TBS
10. The slides are counter-stained for 2 minutes in haematoxylin
11. The slides are then rinsed with de-ionised water
12. Tacha’s bluing solution is applied for 1 minute
13. The slides are rinsed with de-ionised water.

## Notes

### Competing Interest Statement

The authors have declared no competing interest.

### Funding Statement

This study did not receive any funding.

### Author Declarations

Ethics committee/IRB of National Hospital, Abuja gave ethical approval for this work.

